# Microbiota long-term dynamics and prediction of acute graft-versus-host-disease in pediatric allogeneic stem cell transplantation

**DOI:** 10.1101/2021.02.19.21252040

**Authors:** Anna Cäcilia Ingham, Katrine Kielsen, Hanne Mordhorst, Marianne Ifversen, Frank M. Aarestrup, Klaus Gottlob Müller, Sünje Johanna Pamp

## Abstract

**Background:** Patients undergoing allogeneic hematopoietic stem cell transplantation (HSCT) exhibit changes in their gut microbiota and are experiencing a range of complications, including acute graft-versus-host disease (aGvHD). It is unknown if, when, and under which conditions a re-establishment of microbial and immunological homeostasis occurs. It is also unclear whether microbiota long-term dynamics occur at other body sites than the gut such as the mouth or nose. Moreover, it is not known whether the patients’ microbiota prior to HSCT holds clues to whether the patient would suffer from severe complications subsequent to HSCT. Here, we performed integrated host-microbiota analyses of the gut, oral, and nasal microbiotas in 29 children undergoing allo-HSCT.

**Results:** The bacterial diversity decreased in the gut, nose, and mouth during the first month and reconstituted again 1-3 months after allo-HSCT. The microbial community composition traversed three phases over one year. Distinct taxa discriminated the microbiota temporally at all three body sides, including *Enterococcus* spp., *Lactobacillus* spp., and *Blautia* spp. in the gut. Of note, certain microbial taxa appeared already changed in the patients prior to allo-HSCT as compared to healthy children. Acute GvHD occurring after allo-HSCT could be predicted from the microbiota composition at all three body sites prior to HSCT, in particular from *Parabacteroides distasonis*, *Lachnospiraceae* NK4A136 sp. and *Lactobacillus* sp. abundances in the gut. The reconstitution of CD4+ T cells, T_H_17 and B cells was associated with distinct taxa of the gut, oral, and nasal microbiota.

**Conclusions:** This study reveals for the first time bacteria in the mouth and nose that may predict aGvHD. Surveillance of the microbiota at different body sites in HSCT may be of prognostic value and could assist in guiding personalized treatment strategies. The identification of distinct bacteria that have a potential to predict post-transplant aGvHD might provide opportunities for an improved preventive clinical management, including a modulation of microbiomes. The host-microbiota associations shared between several body sites might also support an implementation of more feasible oral and nasal swab sampling-based analyses. Altogether, the findings suggest that both, host factors and the microbiota, could provide actionable information to guiding precision medicine.

## Background

In allogeneic hematopoietic stem cell transplantation (allo-HSCT), the infusion of donor derived stem cells is employed as a curative treatment for various types of hematologic and non-hematologic disorders [1]. In allo-HSCT patients, the human gut microbiota changes subsequent to transplantation, which may in part be attributable to antimicrobial treatment and conditioning regimens [2–4]. Butyrate-producing bacteria affiliated with the order *Clostridiales* are depleted in the gut early after transplantation, while *Proteobacteria*, and *Lactobacillales* such as *Enterococcus* spp. expand, possibly due to both increased oxygen levels in the intestinal lumen in the absence of butyrate, and antimicrobial resistance [2–5]. However, microbiota dynamics in HSCT patients have so far mainly been monitored in detail during the first month post HSCT and not over longer periods of time. Hence, it is unclear whether and when the microbiota re-establishes to similar microbial community structures as prior to HSCT.

Conditioning-induced intestinal epithelial permeability might promote bacterial translocation and bacteremia [6]. This is recognized as the initial step in the pathogenesis of acute graft-versus-host disease (aGvHD) [7]. Acute GvHD is a common side effect of allo-HSCT in which alloreactive donor T cells exhibit cytotoxic activity against healthy tissue in the host, including the gut epithelium [7]. Acute GvHD severity can be distinguished in four grades dependent on the extent of organs affected: Grade 0-I presents as no or mild, and grade II-IV as moderate to severe aGvHD. Recently, studies have suggested that a lower gut microbiota diversity is associated with aGvHD and aGvHD-related mortality and that certain bacterial taxa dominating post HSCT may be involved in promoting aGvHD [3,8–12]. However, it has not been examined whether microbiota composition prior to HSCT has a predictive value in forecasting possible aGvHD severity, and which is addressed in the present study.

The microbiota exerts immunomodulatory function on the host’s adaptive immune system, for example on T cells [13]. For instance, human commensal gut strains affiliated with *Bacteroides* and *Clostridia* can induce T regulatory (T_reg_) cells in germ-free mice [14]. Recent findings suggest that functionally different T cell subsets, such as T helper 17 (T_H_17) and T_reg_ cells are involved in the pathogenesis of aGVHD [15–17]. The microbiota at body sites other than the gut, such as the oral and nasal cavities, have also been suggested to be involved in immunomodulation [18]. We have previously proposed that the gut microbiota is associated with immune cell reconstitution after allo-HSCT [4]. However, it is unknown if the microbiotas at other mucosal sites are affected by allo-HSCT, whether they are associated with aGvHD, and whether they are associated with recovery of the patients’ immune system.

Here, we monitored the microbiota dynamics in the gut, oral, and nasal cavities in pediatric allogeneic HSCT patients over a period of one year. At all three body sites, we identify distinct temporal bacterial abundance trajectories. In a machine learning approach, we predict aGvHD severity from pre-transplant microbiotas in the gut, oral, and nasal cavities which may be useful for early preventive managements in the clinical setting. By relating the microbiota composition to immune cell counts, inflammation and infection markers, antibiotic treatment, clinical outcomes, and patients’ baseline parameters, we uncover similarities in host-microbial associations at different body sites.

## Results

We characterized long-term microbiota dynamics in pediatric allo-HSCT at three body sites: the gut, and oral and nasal cavities (Figure 1). Fecal samples, buccal swabs, and anterior naris swabs were collected from 29 children at 10 time points over a one-year period: Twice prior to HSCT, on the day of HSCT, weekly during the first month after HSCT, and at three follow-up time points up to twelve months post HSCT (Figure 1). Microbial community dynamics in these samples were determined by 16S rRNA gene profiling. A total of 709 patient samples (212 fecal samples, 248 oral swabs, and 249 nasal swabs from 10 time points) were characterized. Upon sequence filtering (see Methods), we retained 2465 ASVs for the fecal, 377 ASVs for the oral, and 197 ASVs for the nasal core microbiota sets. We predicted the development of aGvHD severity from pre-transplant gut, oral, and nasal microbial abundances using machine learning. In addition, we assessed multivariate associations between the microbiota at the different body sites and immune reconstitution, immune markers, and clinical outcomes. Immune reconstitution was determined through quantitative measurements of T, B, and NK cells, and other leukocyte subpopulations in peripheral blood (Figure 1). We assessed systemic inflammation through levels of C-reactive protein (CRP), and measured procalcitonin as an approximation of infection (Figure 1, see Methods).

**Figure 1.**
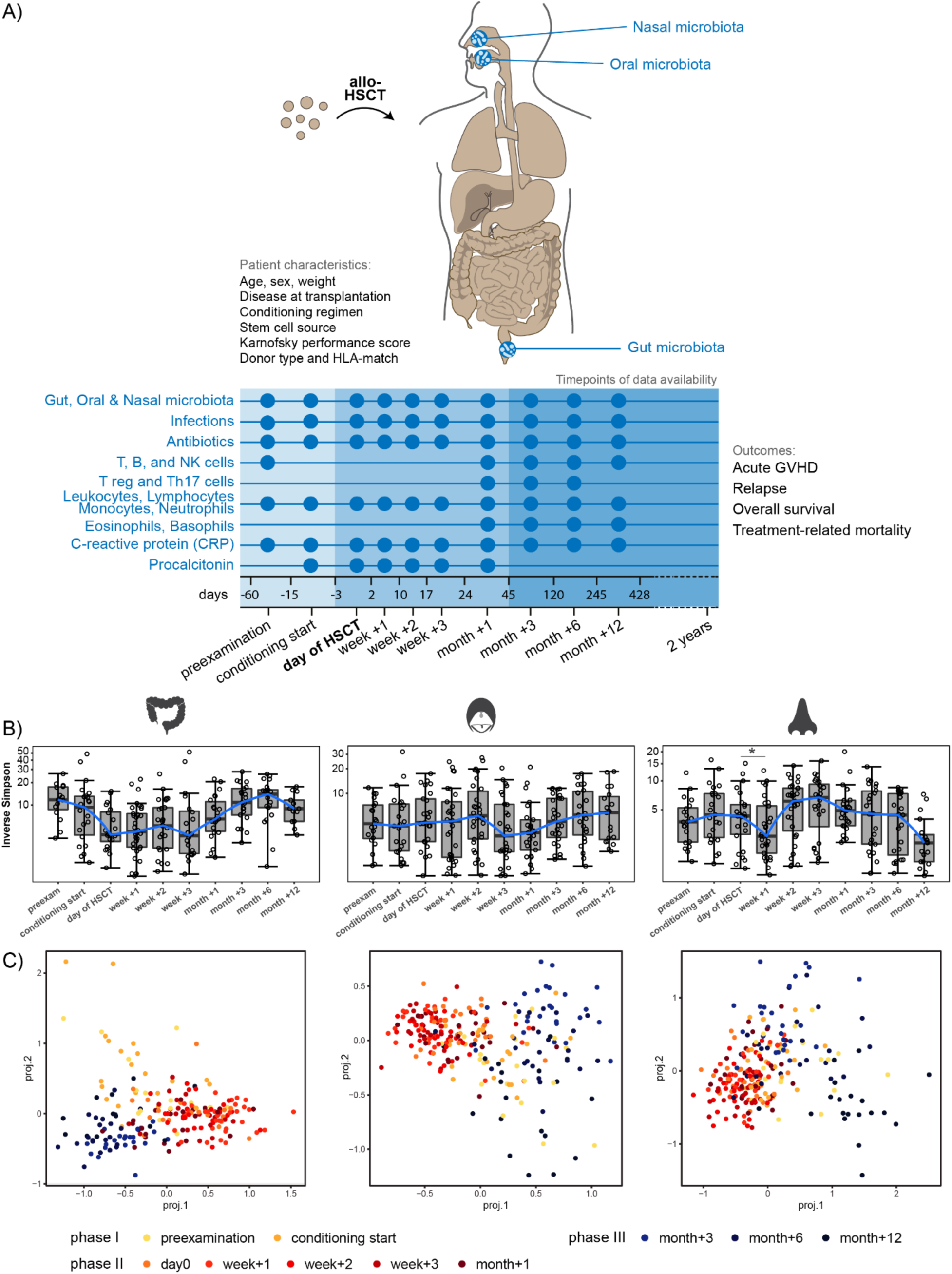
Monitoring gut, oral, and nasal microbiota and the host immune system in allogeneic hematopoietic stem cell transplantation (HSCT). A) Twenty-nine children were monitored before, at the time of, and immediately post allogeneic HSCT, as well as at late follow-up time points. Patients’ baseline characteristics, clinical outcomes, as well as immune cell counts, and inflammation and infection markers over time were monitored. Patient characteristics are described in detail in Table S1 (Additional File 1). Host immune system parameters were related to longitudinal dynamics of the gut, oral, and nasal microbiota that was assessed at the denoted time points. B) Bacterial alpha diversity before, at the time of, and after HSCT at each body site, displayed on a log10 transformed y-axis for visualization purposes. Asterisks indicate significant differences in median inverse Simpson index between time points * *P* < 0.05. C) Tree-based sparse linear discriminant (LDA) analyses by time point in relation to HSCT. For fecal samples, the positive LDA scores were observed for samples collected immediately post HSCT. For both oral and nasal samples, the positive LDA scores were observed for samples from before HSCT and from late follow up-time points.

### Patient cohort and outcomes

The 29 children had a median age of 8.2 years (range: 2.5-16.4) at the time of HSCT. Nine patients (31%) had no or mild aGvHD (grade 0 or I) and 20 patients (69%) developed moderate to severe aGvHD (grade II-IV) at median +14 days following HSCT (range: day +9 to day +61) (Supplementary Table S1, Additional file 1; and https://doi.org/10.6084/m9.figshare.13567502). The main organs involved in aGvHD included the skin (all), intestinal tract (n=3), and the liver (n=2). During the follow-up period of 21.4 months on average (range: 10.1 – 32.7 months), two patients (7%) relapsed and one patient underwent a donor lymphocyte infusion. Three patients (10%) died (one relapse-related death at day +91 and two treatment-related deaths at days +111 and +241, respectively). Due to their low incidence, we did not focus our analysis on relapse and mortality. For 25 patients (86.2%) ≥1 bacterial infection indicated by positive microbial culture was reported throughout the monitored period. All patients were treated prophylactically with trimethoprim and sulfamethoxazole prior to HSCT. In cases of fever or clinical signs of infections, antibiotic treatment with meropenem (28 patients), vancomycin (24 patients), ciprofloxacin (20 patients), phenoxymethylpenicillin (14 patients), or other antibiotics was commenced according to culture-based results or clinical presentation.

### Bacterial alpha diversity decreases in relation to allo-HSCT at all three body sites

Alpha diversity (Inverse Simpson) in the gut was overall the highest, followed by the oral cavity, and the nose (Figure 1B). The lowest alpha diversity was observed within the first month post HSCT for all three body sites. However, the exact time points were somewhat different for each body site: the day of HSCT to week +3 for the gut, week +3 for the oral cavity, and week +1 for the nasal cavity. The decrease in microbial diversity was significant for the nasal cavity, where the median alpha diversity decreased from 4.43 at the start of conditioning to 2.65 in week +1 (*P* = 0.02) (Figure 1B). Alpha diversity increased again at all body sites thereafter. However, alpha diversity was lower again at month +12 in the nasal cavity.

### Microbial community composition in patients prior to HSCT differs from healthy controls

We hypothesized that the bacterial alpha diversity at the first sampling time point (preexamination) might already be lower in these patients as compared to age-matched healthy children due to the treatment given prior to the referral to allo-HSCT and enrolment in this study. To assess this, we compared the gut microbiota at preexamination to that of healthy children (median age 6.8 years) [19]. As expected, the alpha diversity was 2.4-fold lower in the patients at preexamination (median InvSimpson 11.7) as compared to the healthy children (median InvSimpson 28.2) (Supplementary Figure S1A, Additional File 2). Bacterial composition differed between the two groups (anosim, p=0.001, R=0.44, Figure S1B). This difference was to a certain extent due to a larger variation within the HSCT group (betadisper, p<0.001) (Supplementary Figure S1 B, Additional File 2). Through linear discriminant analysis (LEfSe) and differential abundance analysis (DeSeq2), we found taxa that were significantly more abundant in the patients already at preexamination as compared to the healthy controls: these included *Bacilli* (e.g. *Lactobacillus, Enterococcus*), *Erysipelotrichaceae*, and *Enterobacteriaceae* (e.g. *Klebsiella*). In contrast, certain taxa were more abundant in the healthy children, such as *Prevotella*, *Ruminococcaceae* (e.g. *Ruminococcus*), and *Akkermansia*, as compared to the patients at preexamination (Supplementary Figure S1 C and D, Additional File 2; and https://doi.org/10.6084/m9.figshare.13614230).

### Temporal microbial community dynamics appear in three interlaced phases over one year

For a more detailed assessment of gut, oral, and nasal ASVs that best characterized samples from different time points, we performed tree-based sparse linear discriminant analyses (LDA). We observed at all three body sites that samples divided into three partly interlaced phases; phase I: samples at pre-examination and conditioning start, phase II: day of HSCT to month +1, and phase III: month +3 to month +12 (Figure 1C). Interestingly, samples from phase I and III overlapped for the oral and nasal cavities, suggesting a possible return of microbial communities from later time points to a state similar to before HSCT. Of note, the nasal community composition at month +12, that exhibited low alpha diversity, was different from samples of week +1 (phase II) that also exhibited low alpha diversity (Figure 1B and C).

To get a more detailed view of the microbial abundance dynamics, we examined the 12 most abundant families at each body site, respectively (Figures 2A, 3A, Additional File 2: Figures S2 and S3A). In the gut, we observed a reduction in *Lachnospiraceae* in phase II, immediately after HSCT, from 13% at pre-examination to 4.7% in week +1, followed by a recovery to 27.5% in month +3 at the start of phase III (Figure 2A). Concurrently, an expansion of *Enterococcaceae* in phase II (pre-examination: 6.1%; week +1: 22.8%) and *Lactobacillaceae* in phase II (pre-examination: 2%; week +1: 7%) occurred, followed by a reduction in phase III from month +3 onwards to 0.2% and 0.6%, respectively (Figure 2A).

**Figure 2.**
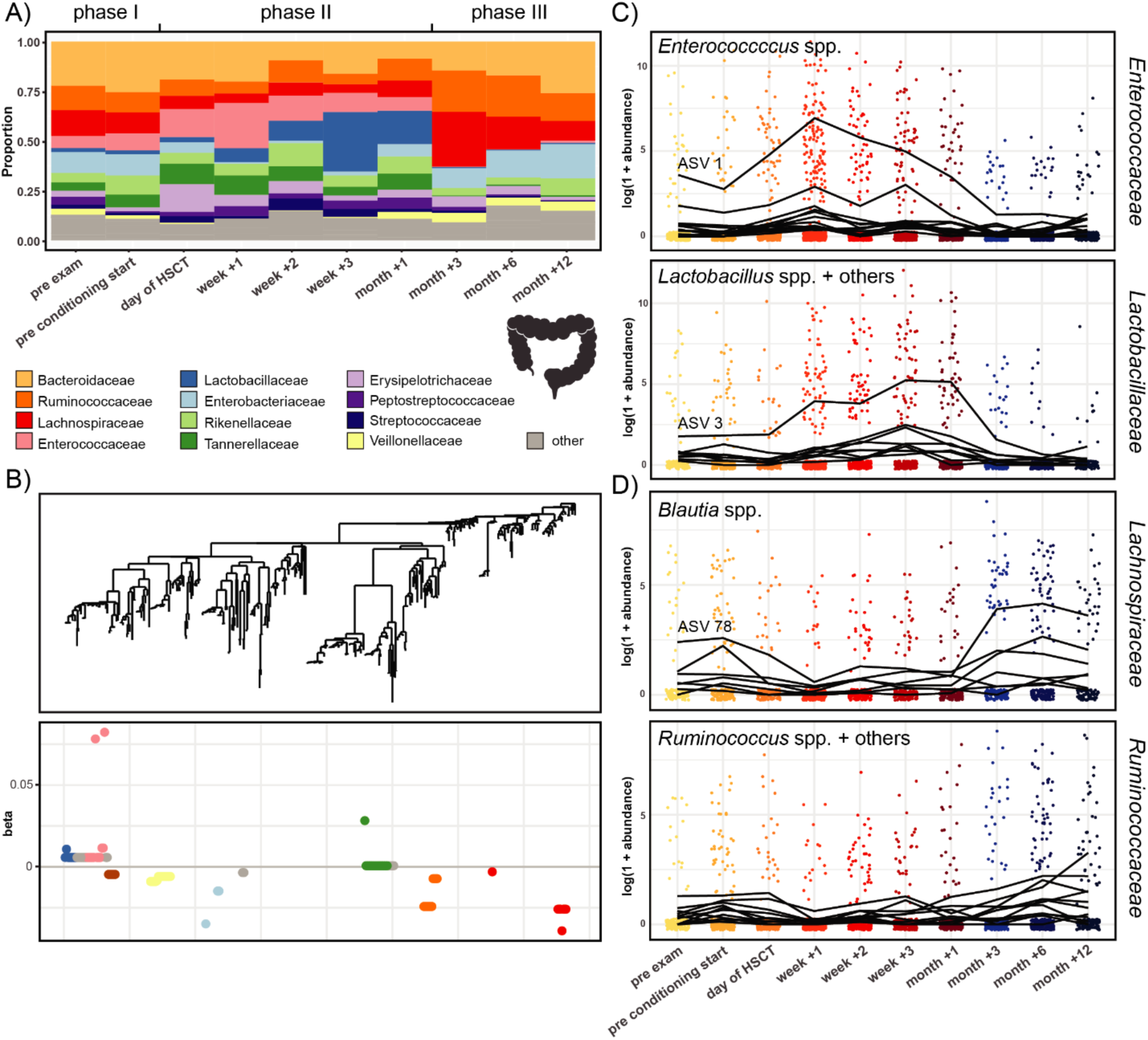
Temporal microbial community dynamics in the gut. A) Relative abundances over time of the 12 most abundant families in the gut. B) Tree-based sparse linear discriminant analysis (LDA). Coefficients of discriminating clades of ASVs on the first LDA axis, colored by taxonomic family, and plotted along the phylogenetic tree. C) Trajectories of ASVs affiliated with the families *Enterococcaceae* and *Lactobacillaceae,* with increasing abundances after HSCT. The most abundant discriminating ASV for each family is indicated. D) Trajectories of ASVs affiliated with the families *Lachnospiraceae* and *Ruminococcaceae*, with decreasing abundances after HSCT and recovery at late follow-up time points. The most abundant discriminating ASV for *Blautia* spp. is indicated. Detailed taxonomic information and LDA-coefficients of the displayed ASVs are listed in Additional File 1: Table S2.

In the oral cavity, we observed a reduced relative abundance of *Actinomycetaceae* for several time points in phase II as compared to the time points in phase I (prior to HSCT) and at later follow-up time points. For example, *Actinomycetaceae* abundances were 9.7% at pre-examination and 2.9% in week +3 (Figure 3A). Furthermore, *Streptococcaceae* abundances were lower from the day of HSCT until week +2 compared with before HSCT and late follow-up time points (pre-examination: 44.6%; week +1: 23.3%; month +3: 51.3%, Figure 3A).

**Figure 3.**
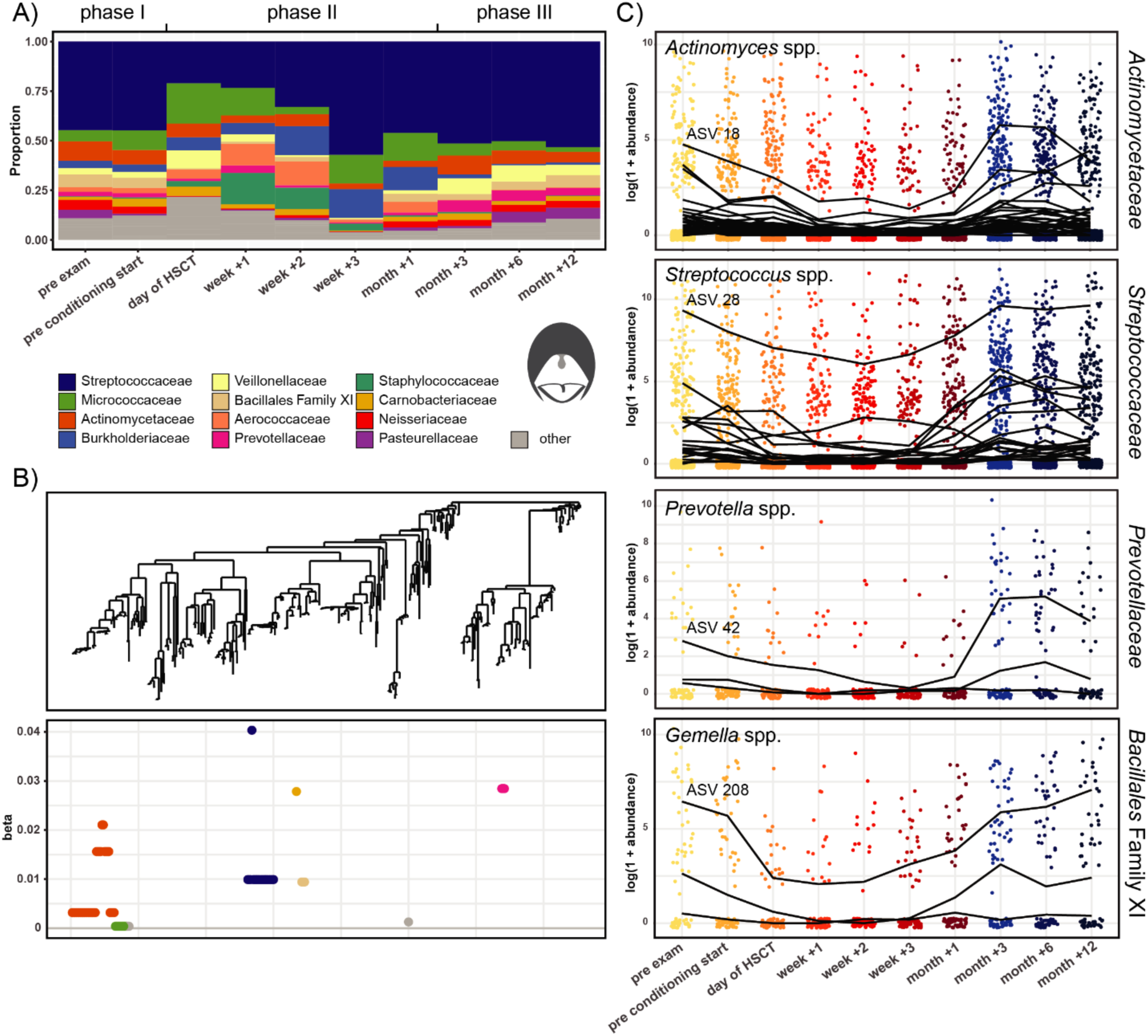
Temporal microbial community dynamics in the oral cavity. A) Relative abundances over time of the 12 most abundant families in the oral cavity. B) Tree-based sparse linear discriminant analysis (LDA). Coefficients of discriminating clades of ASVs on the first LDA axis, colored by taxonomic family, and plotted along the phylogenetic tree. C) Trajectories of ASVs affiliated with the families *Actinomycetaceae, Streptococcaceae, Prevotellaceae, and* Family XI (Class *Bacillales),* with decreasing abundances after HSCT and recovery at late follow-up time points. The most abundant discriminating ASV for each family is indicated. Detailed taxonomic information and LDA-coefficients of the displayed ASVs are listed in Additional File 1: Table S2.

In the nasal cavity, we observed a reduced relative abundance of *Corynebacteriaceae* and *Moraxellaceae* at most time points in phase II, as compared to samples from phase I and III (Additional File 2: Figure S3). For example, *Corynebacteriaceae* abundances were 28.7% at pre-examination and 0.7% in week +1 (Additional File 2: Figure S3).

### Distinct *Enterococcus*, *Lactobacillus*, and *Blautia* lineages discriminate the gut microbiota temporally

In order to determine which specific taxa in the gut were driving the differences between samples in the LDA (Figure 1C), we examined the individual discriminating ASVs. In general, in tree-based sparse LDA, ASVs with positive LDA coefficients are overrepresented in samples with positive LDA scores, while ASVs with negative LDA coefficients likewise are associated with samples with negative LDA scores (Figures 1C, 2B, 2C, and 2D). The LDA revealed 19 clades (total 102 ASVs) in the gut that best separated samples by time point (Figure 2B). The two most discriminating clades with positive LDA-coefficients comprised ASVs of the family *Enterococcaceae* and *Lactobacillaceae* (Figure 2B). The ASVs of these two clades increased in abundance from the day of HSCT (*Enterococcaceae*) and week +1 (*Lactobacillaceae*), respectively, in support of the family abundances and in line with the positive LDA scores of phase II samples (Figures 2A, 2C, and 1C). Of note, the order *Lactobacillales* and genus *Lactobacillus* (family *Lactobacillaceae*) appeared already to be higher at pre-examination as compared to healthy children (Supplementary Figure S1D, Additional File 2). From month +3 onwards, their abundances decreased again to levels comparable to the time of pre-examination (i.e. pre-treatment) (Figure 2C). All members of the *Enterococcaceae* clade, with the exception of one ASV, were *Enterococcus* spp. (Additional File 1: Table S2). The most abundant and most frequently observed *Enterococcus* was ASV 1 (Figure 2C and Additional File 1: Table S2). More detailed sequence analysis of the partial 16S rRNA gene sequence using SINA and BLAST alignments revealed that it belonged to the *Enteroccoccus faecium* group. The most abundant and most frequently observed *Lactococcus* was ASV 3 (Figure 2C and Additional File 1: Table S2), and its partial 16S rRNA gene sequence exhibited a high sequence similarity to *Lactobacillus rhamnosus*.

The two most discriminative clades with negative LDA-coefficients included two individual ASVs and one clade of the *Lachnospiraceae* family, and two *Ruminococcaceae* clades (Figure 2B, Additional File 1: Table S2). The abundances of these ASVs decreased in week +1 and recovered from month +3 onwards, returning to abundances comparable with before HSCT or higher (Figure 2D), in agreement with the abundance patterns for those families (Figure 2A). Of note, the family *Ruminococcaceae* appears already to be lower at pre-examination as compared to healthy children (Supplementary Figure S1 C and D, Additional File 2). All ASVs within the *Lachnospiraceae* group belonged to the genus *Blautia* (Additional File 1: Table S2). The most abundant and most frequently observed *Blautia* was ASV 78 (Figure 2D and Additional File 1: Table S2), and its partial 16S rRNA gene sequence exhibited a high sequence similarity to *Blautia wexlerae*.

### Distinct *Actinomyces* and *Streptococcus* lineages discriminate the oral microbiota temporally

The tree-based sparse LDA identified 10 clades of in total 71 ASVs in the oral cavity that best separated samples by time points along the first axis (Figure 3B). The two largest discriminating groups of ASVs were affiliated with *Actinomycetaceae* and *Streptococcaceae* (Figure 3B, Additional File 1: Table S2). The most abundant and among the most frequently observed ASVs were *Actinomyces* ASV 18 *and Streptococcus* ASV 28 (Figure 3C and Additional File 1: Table S2), and their partial 16S rRNA gene sequence exhibited a high sequence similarity to the *Actinomyces viscosis* and *Streptococcus mitis* groups, respectively. Additional discriminating ASVs were affiliated with *Prevotellaceae*, and *Bacillales* Family XI (*Gemella* spp.), respectively. The most abundant and frequently observed ASVs were affiliated with *Prevotella melaninogenica* (ASV 42) and *Gemella sanguis* (ASV 208). In agreement with the relative family abundance dynamics, these clades shared a pattern of depletion from the day of HSCT or week +1 onwards (phase II), until their abundances recovered from month +3 onwards (phase III) (Figures 3A and 3C) to an abundance similar to before HSCT, as observed for *Ruminococcaceae* and *Lachnospiraceae* in the gut.

### Distinct *Corynebacteriaceae* and *Streptococcaceae* lineages discriminate the nasal microbiota temporally

The LDA revealed 30 discriminating nasal clades on axis 1 (comprising in total 36 ASVs), many of which consisted of individual ASVs (Additional File 2: Figure S3B). ASVs affiliated with the same family did not always covary in abundance. The *Corynebacteriaceae*, *Streptococcaceae,* and *Moraxellaceae* ASVs all had positive LDA-coefficients, i.e. their abundances decreased after HSCT and increased again from month+3 onwards (Additional File 2: Figures S3B and S3C). The most abundant and most frequently observed *Corynebacteriaceae* was ASV 14 (Additional File 2: Figure S3C and Additional File 1: Table S2), and its partial 16S rRNA gene sequence exhibited a high sequence similarity to *Corynebacterium propinquum*.

### Acute GvHD severity can be predicted from gut microbiota composition prior to HSCT

To reveal potential associations between the gut microbiota and the severity of acute GvHD, we examined the 12 most abundant families at each body site in patients with no or mild (grade 0-I) and moderate to severe (grade II-IV) aGvHD. In the gut, *Tannerellaceae* were less abundant at time points before HSCT in patients with grade 0-I compared to grade II-IV, especially at pre-examination and at start of conditioning (Figure 4A). In order to predict aGvHD (grade 0-I versus grade II-IV) from microbial abundances at time points up until the time of stem cell infusion, we implemented machine-learning models (see Methods –Statistical anlysis). This analysis revealed 3 significant predictive ASVs in the gut: ASV 128 (*Parabacteroides distasonis*, Tannerellaceae, *P* < 0.01), ASV 268 (*Lachnospiraceae* NK4A136 group sp., Lachnospiraceae, *P* = 0.01) and ASV 3 (*Lactobacillus* sp., Lactobacillaceae, *P* < 0.01) (Figures 4B and 4C, and Additional File 1: Table S3). This means, high abundances of these ASVs before HSCT were associated with the subsequent development of aGvHD grade II-IV post HSCT (Figure 4C). For instance, all pre-transplant samples with a variance stabilized abundance >5.7 of ASV 128 (*Parabacteroides distasonis*) and 67% with a variance stabilized abundance >3 of ASV 3 (*Lactobacillus* sp.) originated from patients who later developed aGvHD grade II-IV (Figure 4C). In agreement, log transformed relative abundances of these ASVs were mostly higher at pre-examination, conditioning start, and the day of HSCT in patients who later developed aGvHD grade II-IV compared with those exhibiting grade 0-I (Figure 4D). For instance, the average abundance of ASV 128 (*Parabacteroides distasonis*) was 5.5 times higher at pre-examinantion in grade II-IV versus in grade 0-I patients (Figure 4D). The temporal trajectory of ASV 3 (*Lactobacillus* sp.) also revealed a higher abundance at time points up to the transplantation in patients with grade II-IV aGvHD compared to those with grade 0-I (Figure 4E). Within the *Lactobacillaceae* identified by the LDA, this pattern seemed to be restricted to ASV3 (Figure 4E). ASV 128 (*Parabacteroides* distasonis) was part of the discriminating group of *Tannerellaceae* identified in the LDA (Figure 4E, and Additional File 1: Table S3). Its trajectory facetted by aGvHD severity confirmed the observation of increased pre-HSCT abundances in patients with subsequent development of aGvHD grade II-IV (Figure 4E).

**Figure 4.**
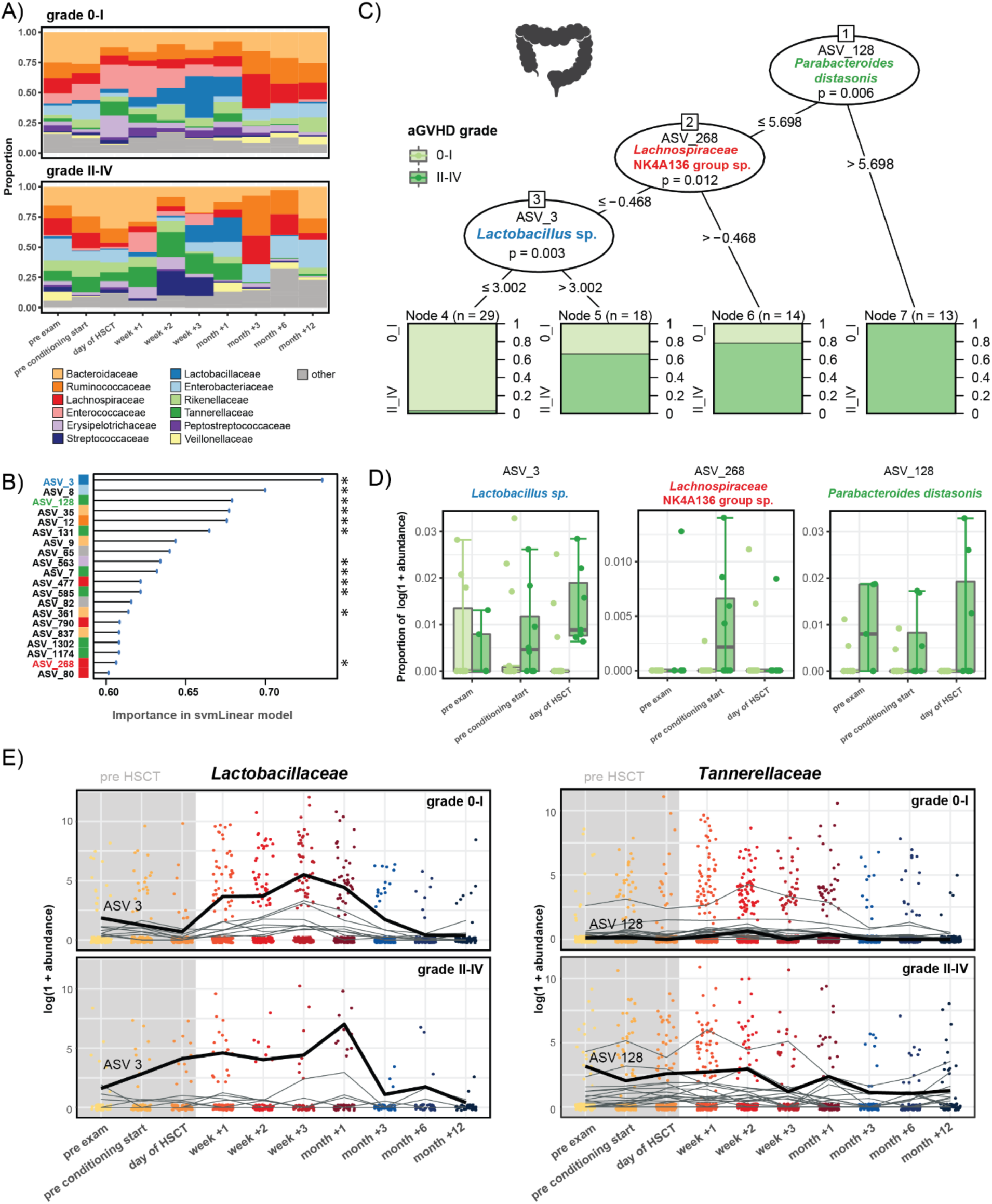
Machine learning-based prediction of aGvHD severity from the pre-HSCT gut microbiota composition. A) Relative abundances of the 12 most abundant families over time in the gut in patients with aGvHD grade 0-I versus II-IV. B) Importance plot of top 20 predictive gut ASVs identified by the svmLinear model with importance scores indicating the mean decrease in prediction accuracy in case the respective ASV would be excluded from the model. The final cross-validated svmLinear model predicted aGvHD (0-I versus II-IV) from the abundances of gut ASVs pre-HSCT with 86% accuracy (95% CI: 65% to 97%). The ASVs that were also confirmed by Boruta feature selection are indicated with asterisk. C) Conditional inference tree (CTREE) displaying ASVs identified as significant split nodes by nonparametric regression for prediction of aGvHD. Numbers along the branches indicate split values of variance stabilized bacterial abundances. The terminal nodes show the proportion of samples originating from patients (n = number of samples) with aGvHD grade 0-I vs II-IV. D) Boxplots depicting the log transformed relative abundances of the predictive ASVs at time points up to the transplantation in aGvHD grade 0-I compared with grade II-IV patients. E) Trajectories of *Lactobacillaceae* and *Tannerellaceae* ASVs that were identified by tree-based sparse LDA, including ASV 3 and ASV 128 that were predictive for aGvHD (bold lines), in patients with aGvHD grade 0-I vs II-IV.

### Acute GvHD severity can be predicted from oral microbiota composition prior to HSCT

In the oral cavity, the bacterial community before HSCT in patients with grade II-IV aGvHD was characterized by a lower relative abundance of *Neisseriaceae*, and higher relative abundances of *Aerococcaceae* and *Prevotellaceae*, compared with grade 0-I aGvHD, especially at pre-examination and conditioning start (Figure 5A). Our machine learning approach predicted aGvHD severity (grade 0-I versus II-IV) from the abundances of 3 significant oral ASVs pre-HSCT: ASV 568 (*Actinomyces* sp., Actinomycetaceae, *P* < 0.001), ASV 226 (*Prevotella melaninogenica*, Prevotellaceae, *P* < 0.001) and ASV 500 (*Pseudopropionibacterium propionicum*, Propionibacteriaceae, *P* < 0.001) (Figures 5B and 5C, and Additional File 1: Table S3). High abundances of these ASVs before transplantation predicted the development of aGvHD grade II-IV after HSCT (Figure 5C). For instance, 91% of samples with a variance stabilized abundance >0.4 of ASV 568 (*Actinomyces* sp.) and 92% of samples with a variance stabilized abundance >6.1 of ASV 226 (*Prevotella melaninogenica*) originated from patients with subsequent development of aGvHD grade II-IV (Figure 5C). In support, pre-HSCT log transformed relative abundances of these ASVs were higher in those patients. For example, the median relative abundance of ASV 500 (*Pseudopropionibacterium propionicum*) on the day of HSCT was 10 times higher in grade II-IV versus in grade 0-I patients (Figure 5D). Temporal trajectories of oral *Actinomycetaceae* and *Prevotellaceae,* identified also in the LDA, showed that the abundances of ASV 226 (*Prevotella melaninogenica*) and ASV 568 (*Actinomyces* sp.) were higher at time points up to the transplantation in patients with grade II-IV versus those with grade 0-I (Figure 5E).

**Figure 5.**
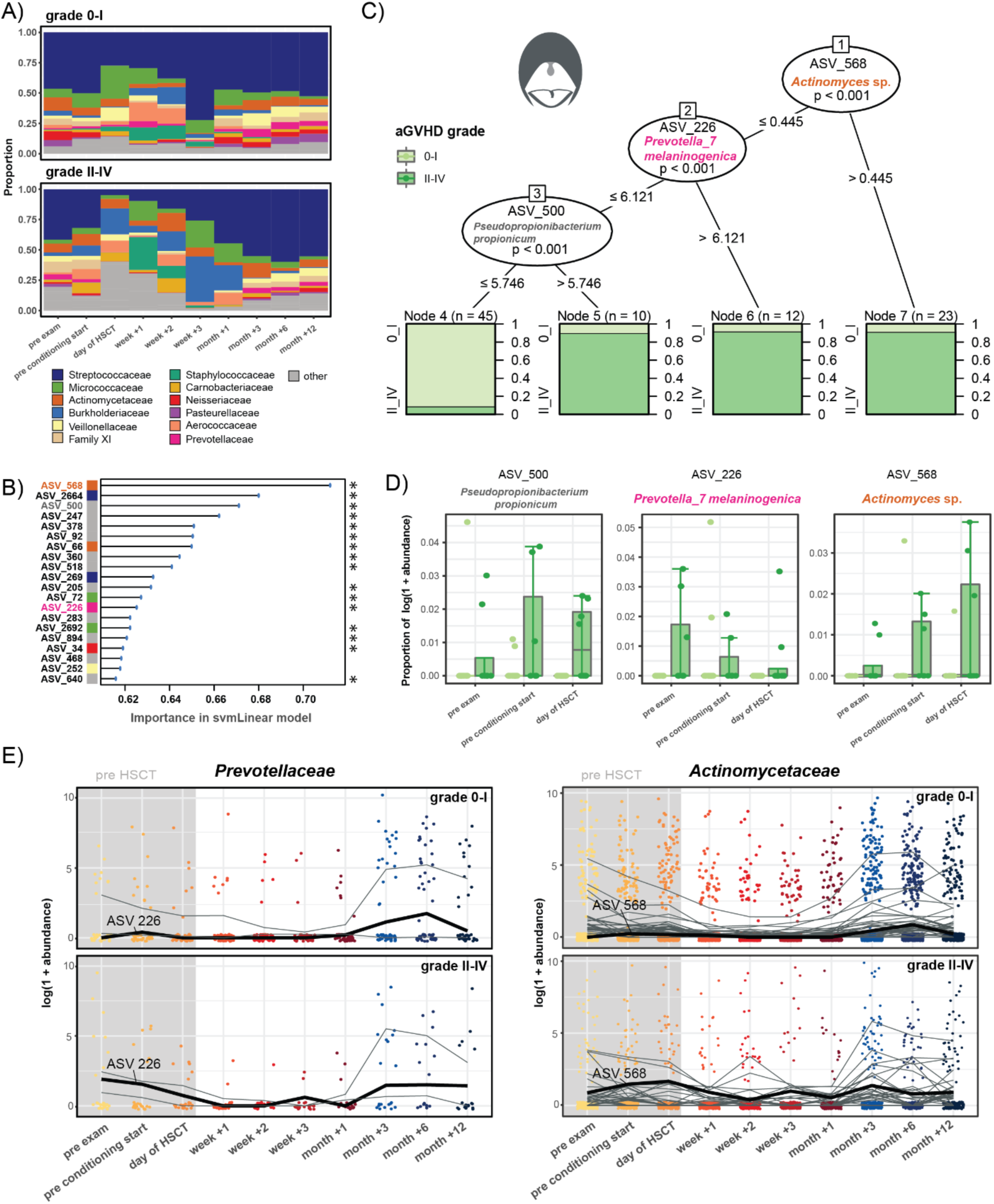
Machine learning-based prediction of aGvHD severity from the pre-HSCT oral microbiota composition. A) Relative abundances the 12 most abundant families over time in the oral cavity in patients with aGvHD grade 0-I versus II-IV. B) Importance plot of top 20 predictive oral ASVs identified by the svmLinear model with importance scores indicating the mean decrease in prediction accuracy in case the respective ASV would be excluded from the model. The final cross-validated svmLinear model predicted aGvHD (0-I versus II-IV) from the abundances of oral ASVs pre-HSCT with 92% accuracy (95% CI: 73% to 99%). The ASVs that were also confirmed by Boruta feature selection are indicated with asterisk. C) Conditional inference tree (CTREE) displaying ASVs identified as significant split nodes by nonparametric regression for prediction of aGvHD. Numbers along the branches indicate split values of variance stabilized bacterial abundances. The terminal nodes show the proportion of samples originating from patients (n = number of represented samples) with aGvHD grade 0-I vs II-IV. D) Boxplots depict the log transformed relative abundances of the predictive ASVs at time points up to the transplantation in aGvHD grade 0-I compared with grade II-IV patients. E) Trajectories of *Prevotellaceae* and *Actinomycetaceae* ASVs that were identified by tree-based sparse LDA, including ASV 226 and ASV 568 that were predictive for aGvHD (bold lines), in patients with aGvHD grade 0-I vs II-IV.

### Acute GvHD severity can be predicted from nasal microbiota composition prior to HSCT

The proportion of nasal *Neisseriaceae* prior to HSCT was higher in patients with aGvHD grade 0-I as compared to grade II-IV (Additional File 2: Figure S4A). In contrast, *Actinomycetaceae* and *Corynebacteriaceae* exhibited a higher abundance in aGvHD grade II-IV patients prior to HSCT compared to those with grade 0-I (Additional File 2: Figure S4A). We found two ASVs significantly predicting aGvHD grade with opposite effects, ASV 66 and ASV 47. A high pre-HSCT abundance of ASV 66 (*Actinomyces* sp., *Actinomycetaceae*, *P* = 0.03) predicted development of aGvHD grade II-IV. The partial 16S rRNA gene sequence of ASV 66 exhibited a high sequence similarity to *Actinomyces viscosus*. A total of 94% of samples with a variance stabilized abundance >6.4 of ASV 66 originated from patients with subsequent development of aGvHD grade II-IV (Additional File 2: Figures S4B and S4C). In support, pre-HSCT log transformed relative abundances of ASV 66 (*Actinomyces* sp.) were 2.3 times higher in patients with aGvHD grade II-IV compared to those with grade 0-I (Additional File 2: Figure S4C). In contrast, high pre-HSCT abundance of ASV 47 (*Rothia* sp., *P* = 0.03) predicted that patients would be spared from aGvHD. The partial 16S rRNA gene sequence of ASV 47 exhibited a high sequence similarity to *Rothia aeria*. All nasal samples with a variance stabilized pre-HSCT abundance >-3.05 of ASV 47 (*Rothia* sp.) originated from patients who subsequently developed no or mild aGvHD (grade 0-I) (Additional File 2: Figure S4B and S3C).

### Reconstitution of CD4+ T cells and the T_H_17 subpopulation is associated with gut, oral, and nasal microbiota

In order to characterize associations between the microbiota and immune cell counts, immune markers, and clinical outcomes in HSCT that potentially might impact our predictions of aGvHD, we implemented two multivariate multi-table approaches, namely sparse partial least squares (sPLS) regression and canonical correspondence analyses (CCpnA). Using sPLS regression, we identified three clusters of ASVs for each body site, respectively (Figures 6A, and Additional File 2: S5A and S6A), which was supported by the CCpnA (Figure 6B, and Additional File 2: S5B and S6B). Several cell populations of the adaptive immune response were associated with one cluster each at all three body sites according to the sPLS analysis. These included T cell counts at late follow-up time points, particularly CD4+ T cells in months +3 and +6, and the subpopulation of T_H_17 cells in months +1 and +3. In the gut, high numbers of these adaptive immune cell populations were associated with high abundances of mainly *Lachnospiraceae*, *Ruminococcaceae*, and *Lactobacillaceae* ASVs (gut cluster 1, Figure 6A). Of note, two of the *Lactobacillus* spp. ASVs in gut cluster 1 (ASV 31 and ASV 586) were also observed as members of the group of *Lactobacillaceae* that discriminated samples from different time points in the LDA (Figure 2C). In the oral cavity, the same lymphocyte subsets were positively correlated with specific *Flavobacteriaceae*, *Prevotellaceae*, *Veillonellaceae*, and *Neisseriaceae* ASVs (oral cluster 3, Additional File 2: Figure S5A). The nasal cluster 1 that was affiliated with high T cell counts comprised predominantly *Veillonellaceae* (Additional File 2: Figure S5A). The nasal cluster 3 was characterized by high T cell counts at pre-examination and exhibited a high abundance of ASV 47 (*Rothia* sp.) and other taxa that were associated with no to mild aGvHD (grade 0-I) (Additional File 2: Figure S4).

**Figure 6.**
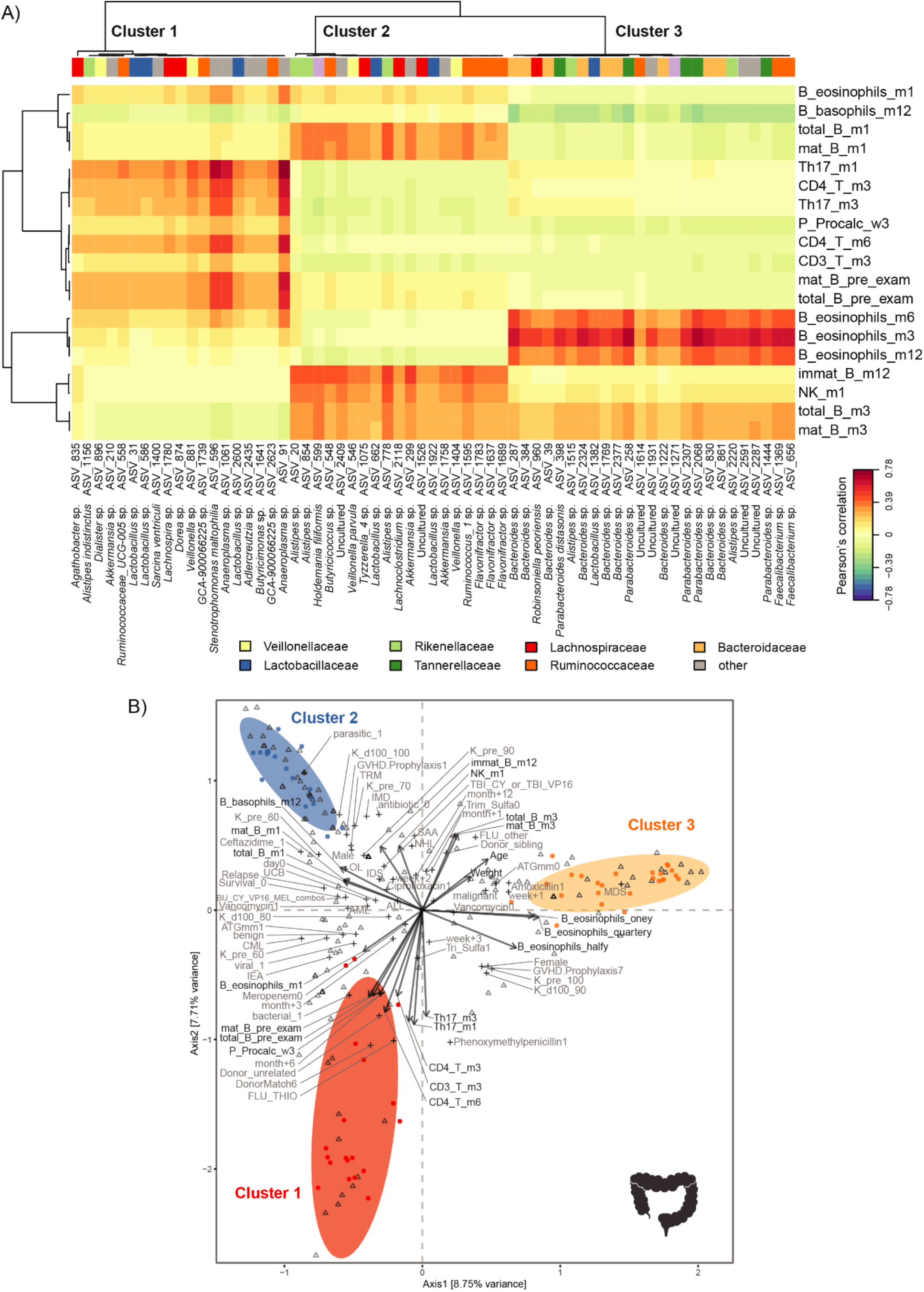
Multivariate associations of the gut microbiota with immune and clinical parameters in HSCT. A) Clustered image map (CIM) based on sparse partial least squares (sPLS) regression analysis (dimensions 1, 2, and 3) displaying pairwise correlations >0.3/<-0.3 between ASVs (bottom) and continuous immune and clinical parameters (right). Red indicates a positive correlation, and blue indicates a negative correlation, respectively. Based on the sPLS regression model, hierarchical clustering (clustering method: complete linkage, distance method: Pearson’s correlation) was performed resulting in the three depicted clusters. B) Canonical correspondence analysis (CCpnA) relating gut microbial abundances (circles) to continuous (arrows) and categorical (+) immune and clinical parameters. ASVs and variables with at least one correlation >0.3/<-0.3 in the sPLS analysis were included in the CCpnA. The triplot shows variables and ASVs with a score >0.3/<-0.3 on at least one of the first three CCpnA axes, displayed on axis 1 versus 2 with samples depicted as triangles. The colored ellipses (depicted with 80% confidence interval) correspond to the clusters of ASVs identified by the sPLS-based hierarchical clustering. Abbreviations not mentioned in text: ATGmm, anti-thymocyte globulin; B_, blood; BU, busulfan; CY, Cyclophosphamide; DonorMatch6, matched unrelated donor; FLU_other, fludarabine combinations without thiotepa; GvHD.Prophylaxis1, treatment with cyclosporine; GvHD.Prophylaxis7, treatment with cyclosporine and methotrexate; immat_B, immature B cells; K_d100, Karnofsky score on day +100; K_pre, Karnofsky score before HSCT; m1, month+1; m3, month+3; m6, month+6; m12, month+12; mat_B, mature B cells; MEL, melphalan; total_B, total B cells; P_, plasma; parasitic, parasitic infection; pre_cond, before conditioning start; pre_exam, pre-examination; THIO, thiotepa; viral, viral infection; VP16, Etoposide.

In the CCpnA, we observed that samples in gut cluster 1 (mainly from months +3 and +6) belonged to patients with benign primary diseases, who received conditioning regimens involving fludarabine (Figure 6B). Moreover, these patients had a high number of bacterial and viral infections and were treated often with phenoxymethylpenicillin compared to the overall patient population. In the oral cavity, samples associated with CD4+ T cell reconstitution similarly stemmed from late follow-up time points and from pre-examination. Patients in oral cluster 3 were generally treated with few antibiotics. The CCpnA of the nasal data set indicated that patients with high CD4+ T cell and T_H_17 cell counts at late follow-up time points exhibited moderate to severe aGvHD (grade II-IV). Furthermore, these patients were treated with meropenem, ciprofloxacin, and vancomycin more often compared with the remaining patient population (Figure S6B). Most samples in the nasal cluster 1 were collected in weeks +2 and +3.

### Reconstitution of B cells is associated with gut, oral, and nasal microbiota

At all three body sites, B cell counts at several late follow-up time points exhibited associations with microbial abundances. High B cell counts were positively correlated with high abundances of *Ruminococcaceae*, *Lachnospiraceae*, and *Rikenellaceae,* as well as few *Veillonellaceae* and *Lactobacillaceae* in the gut (cluster 2, Figure 6A). In addition, the gut cluster 2 was associated with high NK cell counts in month +1. In the oral cavity, ASVs within the small cluster 1, particularly ASV 422 (*Actinomyces odontolyticus*) and ASV 546 (*Veillonella parvula*), were positively correlated with these cell counts, whereas ASVs affiliated with *Staphylococcaceae* and *Lactobacillaceae* (oral cluster 2) exhibited negative correlations (Additional File 2: Figure S5A). ASV 422 (*Actinomyces odontolyticus*) was also observed within the group of *Actinomycetaceae* ASVs in the LDA of the oral microbiota. In the nasal cavity, abundances of *Streptococcaceae*, *Moraxellaceae*, and *Corynebacteriaceae* within nasal cluster 3 were positively correlated with high B cell counts, particularly in month +3 (Additional File 2: Figure S6A). The CCpnA indicated that samples in gut cluster 2 were taken predominantly in week +2, whereas samples in oral cluster 1 were mainly collected in months +3 and +6 (Figures 6B and Additional File 2: Figure S5B). Both the gut and oral CCpnA indicated that the associations between B cell counts and microbial abundances predominantly occurred in patients who underwent a conditioning regimen without TBI and without fludarabine (in contrast to conditioning regimens involving TBI or fludarabine). Furthermore, these patients were treated with ceftazidime, vancomycin, and ciprofloxacin, but sparsely with other antimicrobial agents (Figure 6B and Additional File 2: Figure S5B). The CCpnA on the gut data set revealed that samples in this cluster (gut cluster 2) originated from both patients diagnosed with malignant diseases and benign diseases (Figure 6B).

### Body site-specific immune-microbial associations

In addition to immune-microbial associations shared between two or three of the examined body sites, we observed a few patterns that were exclusive to individual sites. In cluster 3 in the gut, we observed ASVs primarily affiliated with *Bacteroidaceae* and *Tannerellaceae* whose abundances showed positive correlations with eosinophil counts in months +3, +6, and +12. In the oral cavity, the sPLS analysis revealed a sub-cluster of oral cluster 3 comprising ASVs affiliated with various families, e.g. ASV 1172 (*Actinomyces sp.*), which was also identified as one of the discriminating *Actinomycetaceae* ASVs in the LDA. In the sPLS analysis, this sub-cluster was associated with high counts of T_reg_ and T_H_17 cells at late follow-up time points (Additional File 2: Figure S6A).

## Discussion

Both the microbiota and the immune system are subject to major changes during allogeneic HSCT. Failure to re-establish host-microbial homeostasis might have adverse consequences for the patients, such as prolonged immune deficiency. Long-term surveillance of microbial dynamics is required to understand i) the shifts in the microbial community structure induced by HSCT and its accompanying treatments, and ii) at which time points and under which conditions re-establishment of immunological and microbial homeostasis occurs. Such knowledge may be of great prognostic value and may assist in guiding personalized treatment strategies. Here, we present a comprehensive assessment of temporal microbial abundance trajectories from before, at the time of, and after HSCT, to late follow-up time points up to one year.

We have identified a group of *Ruminococcaceae*, and a clade of *Blautia* spp. (*Lachnospiraceae*), temporally discriminating microbial community structure in the gut in relation to HSCT. We show a clear pattern of depletion of fecal *Blautia* spp. immediately post HSCT, as well as their recovery from month +3 post HSCT onwards. One could describe the trajectories of these potentially beneficial taxa as a “smile”-shape. Previous studies have associated the taxonomic families of *Ruminococcaceae* and *Lachnospiraceae* (both class *Clostridia*), and especially the genus *Blautia* (family *Lachnospiraceae*), with lower mortality, lower GvHD, and higher bacterial diversity in adult allo-HSCT recipients [4,9,20–22]. In turn, a loss of those taxa after HSCT was associated with subsequent adverse outcomes. Our findings extend the potential of *Blautia* spp. abundances as an indicator of favorable clinical outcomes, as we characterize abundance dynamics in children and provide important insight into the time point for the expected return to abundances comparable to pre-HSCT time points (i.e. between month +1 and +3).

Adverse effects, like bacteremia and GvHD, have been found to accompany an expansion of the genus *Enterococcus* post transplantation [2,3,6,23]. We have found a characteristic expansion of this genus, as well as of certain *Lactobacillaceae* after HSCT, in agreement with other recent studies [4,6,11]. In addition, we were able to show a decrease of *Enterococcus* spp. and *Lactobacillaceae* from month +3 to abundances comparable to pre-HSCT levels. The abundance of these taxa over the course of one year might be described as a “frown”-shaped trajectory. As for the “smile”-trajectory of potentially beneficial taxa, the “frown”-trajectories of these taxa could be the first step towards a novel basis to evaluate the re-establishment of patients’ microbial homeostasis and associated convalescence. Importantly, *Enterococcus* was already higher abundant in the patient cohort at preexamination prior to HSCT as compared to the healthy age-matched cohort, most likely due to prior chemotherapy and antibiotic treatment given before referral to HSCT. Knowledge about the abundance level of *Enterococcus* before HSCT could therefore provide valuable information about potential high-risk individuals already prior to transplantation. It should be noted, however, that despite the observed different abundance levels in patients and healthy controls, and the further expansion of *Enterococcus* post HSCT being in line with previous studies, our multivariate analyses did not reveal direct detrimental host-microbial associations of *Enterococcus* in the present cohort.

We have to our knowledge for the first time determined long-term dynamics of the oral and nasal microbiota in allogeneic HSCT patients. Interestingly, we identified abundance trajectories of phylogenetically closely related groups of *Actinomycetaceae*, *Streptococcaceae*, *Prevotellaceae*, and Family XI (*Gemella* spp., Class *Bacillales)* in the oral cavity, resembling the “smile”-shaped trajectories observed in gut. These taxa are part of the normal oral microbiota. Our findings are in agreement with previous studies reporting the detection of fewer *Prevotella* spp. and *Streptococcus* spp. in the oral cavity during the first month post HSCT [24, 25]. In addition, our current study provides insight into the time of recovery of these taxa in month +3 after HSCT.

For the oral cavity, a post-transplant expansion of *Enterococcus* spp. and *Staphylococcus* spp. has been reported previously [25, 26]. Consistently, we observed an increased relative abundance of *Staphylococcaceae* during the first month post HSCT, but we did not identify *Enterococcus* spp. or *Staphylococcus* spp. as significant drivers of temporal dynamics in the oral cavity. Previously, increased *Enterococcus* abundances post HSCT were found predominantly in patients who developed oral mucositis, which was not directly assessed in our study [25, 27]. Therefore, our findings suggest that further investigation of taxa that exhibit “smile”-like abundance trajectories could be relevant in direct relation to oral mucositis. Especially *Actinomycetaceae*, *Streptococcaceae*, and *Prevotellaceae*, when low-abundant, might be candidates for bacterial predictors of oral mucositis, and furthermore might be employed to facilitate preventive management.

In the nasal cavity, the microbiota did not exhibit temporal patterns as distinct as the “smile“- and “frown“-shaped trajectories in the gut and the oral cavity. One could speculate that nasal bacterial abundance patterns might be more individualized, which might in turn conceal pronounced patterns when looking at the patient population as a whole. However, certain host-microbial associations observed in the gut were reflected in the nasal cavity. For instance, reconstitution of CD4+ T cells and the T_H_17 subset were associated with distinct groups of ASVs at all three body sites.

Together, these findings suggest that the oral and potentially also the nasal cavity might constitute easily accessible microbial niches suitable for investigating host-microbial associations in the context of HSCT, similar to current strategies for the gut. While mucous membranes that are in close association with distinct microbial communities characterize all three niches, it is more feasible to collect buccal and anterior naris swabs during clinical routine as compared to collecting fecal samples. Fecal sample collection is dependent on bowel movements, which often are impaired in this patient group. Therefore, our study provides valuable knowledge for possible future applications that could include the monitoring of oral microbial dynamics in clinical routine, which might be easier to implement than routine fecal sampling.

We identified AVSs with the potential to predict post-transplant aGvHD, which might open opportunities to improved preventive clinical management, for example by intensified prophylactic immunosuppression for patients at increased risk. Some ASVs were significant for both, discriminating the microbiota in long-term dynamics as well as in the prediction of aGVHD severity from the microbiotas prior to HSCT, such as ASV 3 (*Lactobacillus* sp.) in the gut, as well as ASV 568 (*Actinomyces* sp.) and ASV 226 (*Prevotella melaninogenica*) in the oral cavity. While we do not yet understand the biological mechanisms underlying this observation, these taxa could be of particular interest for a long-term monitoring in pediatric HSCT patients, starting prior to HSCT. Like the gut microbiota, the oral and nasal commensal residents might be of systemic relevance, and a more holistic picture of microbial influences might be drawn by examining various niches with bacterial communities potentially interacting across body sites. In light of intimate host-microbiota interactions, the microbial community patterns might also be a marker for underlying changes occurring in the immune system.

High abundances at late follow-up time points of two fecal *Lactobacillus* spp. that expanded after HSCT showed positive correlations with T cell reconstitution. This is in line with previous studies suggesting that the expansion of *Lactobacillus,* a genus commonly associated with probiotic properties, might promote immune homeostasis and thereby exert a protective effect to limit *Enterococcus* expansion [4,23,28]. A potential explanation indicated by our results might be that high *Lactobacillus* abundances outlasting enterococcal dominance promotes T cell reconstitution. However, the associated cell populations include T_H_17 cells which can facilitate inflammation, and therefore it is difficult to determine whether the observed *Lactobacillus* expansion is exclusively beneficial [13]. However, Th17 cells could perhaps add to the host defense in these patients and therefore be beneficial for local homeostasis, although with the unusual cost of harmful inflammation.

Furthermore, we found associations of high *Lachnospiraceae* and *Ruminococcaceae* in the gut with rapid B and NK cell reconstitution, which is in support of our previous study [4]. These two *Clostridiales* families play an important role in providing the host with short-chain fatty acids (SCFAs), such as butyrate [5, 29]. A study demonstrated that SCFAs can facilitate the differentiation of human naïve B cells to plasma cells in culture [30]. Whether SCFAs also directly influence B cell proliferation is yet unknown.

We have made several observations in which infections and/or antibiotic treatments were associated with the abundance of specific bacterial clusters at certain body sites, immune cell counts, and aGvHD. For example, patients whose samples were represented by gut microbiota cluster 1 experienced a high number of infections and were treated often with phenoxymethylpenicillin compared to the overall patient population. In contrast, patients affiliated with gut microbiota cluster 2 experienced treatment with ceftazidime, vancomycin, and ciprofloxacin, but sparsely with other antimicrobial agents. Furthermore, patients affiliated with oral microbiota cluster 3 were generally treated with few antibiotics, and, patients whose sample were represented by the nasal microbiota cluster 1 were treated often with meropenem, ciprofloxacin, and vancomycin compared with the remaining patient population. However, it is challenging to interpret these observations, as these patient samples were also associated with other features, such as an increased or decreased abundance of certain immune cells (see Additional file 3 for further discussion), or the patients were exposed to other treatments as well, such as TBI or fludarabine. Overall, however, our observations are consistent with previous reports that antimicrobial treatment is associated with changes in microbiota composition in patients undergoing allo-HSCT and might impact clinical outcomes [4,11,31–33]. It will be important to gain a more mechanistic understanding of the possible effects of antimicrobial treatment to disentangle the effect of antibiotics from that of other medications and host responses. Such insight could for example allow selecting more suitable antimicrobials for treatment in HSCT patients that spare the elimination of beneficial taxa, whose decline might be associated with more severe clinical outcomes. The choice of antibiotic treatment might also be important to take into consideration in patients that might potentially be referred to HSCT eventually, given that we already observed certain changes in the microbiota in the patients at referral compared to healthy controls. The microbiota at referral already exhibited some features that were associated with more severe side effects.

Associations between aGvHD severity and the microbiota have to date merely been based on logistic regression and correlation analyses [8,34–36]. In addition, microbial abundances at the time of neutrophil recovery or engraftment were assessed, i.e. at time points shortly before, concurrent to, or potentially after aGvHD onset [8,16,36]. Here, we have implemented machine learning techniques to take the assessment of microbiota-aGvHD relations from correlative to predictive modeling: We presented evidence that aGvHD severity may be predicted from pre-HSCT microbial abundances in the gut, as well as in the oral and nasal cavities. This could open up opportunities for the future where microbial markers guide early interventions to prevent aGvHD. This could include a modulation of the microbiota of patients predicted to be at high risk with synthetic microbiotas containing beneficial bacteria, including probiotics.

Notably, we have to our knowledge for the first time revealed microbial taxa in the oral and nasal cavity that may predict aGvHD. A further discussion on possible connections between specific microbial taxa of the gut, oral, and nasal cavity, immune responses, and aGvHD can be found in Additional file 3.

## Conclusions

With the present study we bring forward a comprehensive framework of host-microbial associations in allogeneic HSCT. We focused on long-term microbial dynamics, demonstrating distinct microbial abundance patterns of disturbance and recovery, as well as making predictions about aGvHD from the pre-transplant microbiota. We discovered that the microbial community composition in patients prior to HSCT already differs somewhat from healthy controls in regard to key microbial taxa, opening up opportunities for potential preventive measure in the future. Moreover, we confirmed the depletion of *Blautia* spp. and expansion of *Enterococcus* spp. in the gut after HSCT and expand this knowledge by precisely defining which phylogenetically closely related sequence variants of these genera are characteristic for those patterns, and when they return to pre-HSCT levels. We identified similar patterns for members of the oral and nasal microbiota and propose month +3 post-transplant as a possible universally crucial time point for microbiota reconstitution after HSCT. We demonstrate that high abundances of for example an intestinal *P. distasonis* ASV, and an oral *P. melaninogenica* ASV pre-HSCT predict the development of moderate to severe aGvHD post-transplant. When relating microbial abundances with immune cell counts, we found rapid B and NK cell reconstitution to be associated with high abundances of *Lachnospiraceae* and *Ruminococcacea*, which also depended on antibiotics treatment. Distinct ASVs at all three body sites were associated with T_H_17 cell counts, suggesting future research on a potential immunomodulatory involvement of the microbiota in inflammation regulation, which might play a role for aGvHD development. We have discovered host-microbial associations shared between two or more of the examined body sites. This may open up opportunities for implementing a more feasible oral and nasal swab sampling into research and clinical diagnostic activities to design more precise patient treatment strategies to reduce serious side effects and improve immune and microbiota reconstitution.

## Materials and Methods

### Patient recruitment and sample collection

We recruited 29 children (age range: 2.5 - 16.4 years) who underwent their first myeloablative allogeneic hematopoietic stem cell transplantation at Copenhagen University Hospital Rigshospitalet (Denmark) between November 2015 and October 2017. We provide detailed information about the patients’ clinical characteristics in Table S1 (Additional File 1). Every patient underwent a myeloablative conditioning regimen starting on day −10 for patients receiving a graft from a haploidentical donor, and on day −7 for patients with sibling or matched unrelated donors (Additional File 1: Table S1). One patient had a donor lymphocyte infusion on day +223 after the first transplantation. Immune cell count date of this patient was excluded from our analysis from the time of donor lymphocyte infusion. We grouped the patients into four categories of conditioning regimens: 1. TBI_CY_or_TBI_VP16 (n=6; TBI + cyclophosphamide or TBI + etoposide), 2. BU_CY_VP16_MEL_combos (n=6; Combinations of busulfan, cyclophosphamide, etoposide and melphalan), 3. FLU_THIO (n=12; subgroups: fludarabine + busulfan + thiotepa (n=6); fludarabine + treosulfan + thiotepa (n=4); fludarabine + thiotepa (n=1); fludarabine + cyclophosphamide + thiotepa (n=1)), and 4. FLU_other (n=5; subgroups: fludarabine + busulfan (n=2); fludarabine + cyclophosphamide (n=2); fludarabine + treosulfan (n=1)) (Additional File 1: Table S1). The following sampling time points were defined: pre-examination (between day −57 and day −15), around the start of conditioning (between day −14 and day −3 and latest 2 days after conditioning start), at time of HSCT (between day −2 and day +2), and weekly during the first 3 weeks after transplantation (week +1: day +3 to day +10, week +2: day +11 to day +17, week +3: day +18 to day +24) (Figure 1A). Broader intervals applied to follow-up time points: Month +1 (between days +25 and +45), month +3 (between days +46 and +120), month +6 (between days +121 and +245), and month +12 (between day +246 and +428). Acute GvHD was graded by daily clinical assessment of skin, liver and gastro-intestinal manifestations according to the Glucksberg criteria [37]. We group aGvHD severity into grade 0-I and grade II-IV, reflecting clinical practice where grade I represents limited alloreactivity with no (or very limited) impact on the overall clinical outcome of HSCT, and therefore no need for medical treatment of these patients, such as the use of glucocorticoids, which is first-line treatment for grade II-IV aGvHD.

To address certain specific questions, we also analyzed the microbiota (from time point 0) of a cohort of 18 healthy children that were part of a previous study [19]. The median age of these children was 6.8 years (interquartile range 4.6 to 9.6). A total of 30 fecal samples were obtained (11 children provided two samples each within an interval of six months). The children did not receive any antibiotics within the month prior to sample collection. The samples were processed in the same way as the fecal samples of the patients of this study (described below).

### Infections and antibiotics

Records of bacterial, fungal, viral, and parasitic infections and antibiotic treatment from before HSCT (from day −30 or at the collection time of the first microbiota sample in case this was earlier) until month +12 (day +428) were taken into consideration (or as long as data was available for the most recent patients; data accessed in July 2018). This corresponds to the length of the sampling period of fecal and swab samples.

### Analysis of immune cell subpopulations

Leukocyte counts were recorded daily during hospitalization starting prior to HSCT, and later weekly in the outpatient clinic by flow cytometry (Sysmex XN) or microscopy (CellaVision DM96 microscope) in case of very low counts. Monitored subpopulations included lymphocytes, monocytes, neutrophils, basophils, and eosinophils.

### Analysis of T, B and NK cells in peripheral blood

T, B, and NK cell counts in x10^9^/L were determined at pre-examination, and in month +1, +3, +6, and +12. Trucount Tubes (Becton Dickinson, Albertslund, Denmark) were used to quantify these cell types in peripheral blood on a FC500 flow cytometer (Beckman Coulter, Copenhagen, Denmark). For immunofluorescence staining, the following conjugated monoclonal antibodies were used for CD3+ T cells, CD3+CD4+ T cells and CD3+CD8+ T cell quantification: CD3-PerCP, CD3-FITC, CD4-FITC, CD8-PE (Becton Dickinson). CD45-PerCP, CD16/56-PE antibodies were used to determine NK cells based on their CD45+CD16+CD56+ phenotype. For B cells, total B cells (CD45+CD19+), mature B cells (CD45+CD19+CD20+) and immature B cells (CD45+CD19+CD20-) were differentiated by using CD20-FITC and CD19-PE antibodies.

### Subtyping of T cells

Peripheral blood samples were collected in month +1, +3 and +6 for isolation of peripheral blood mononuclear cells (PBMCs) by gradient centrifugation of heparinized blood with Lymphoprep™ (Axis-Shield, Oslo, Norway). PBMCs were washed in PBS (Life Technologies, Invitrogen, Paisley, U.K.) three times and then resuspended in RPMI 1640 buffer containing HEPES (Biological Industries Israel Beit-Haemek Ltd, Kibbutz Beit-Haemek, Israel), L-glutamine (GIBCO, Invitrogen, Carlsbad, CA) and Gentamycin (GIBCO), 30% fetal bovine serum (Biological Industries) and 10% Dimethyl Sulfoxide (VWR, Herlev, Denmark) for cryo-preservation in liquid nitrogen.

T cell subsets, i.e. T_H_17 cells and T_reg_ cells, were quantified from frozen PBMCs by flow cytometry on a FACS Fortessa III flow cytometer (Becton Dickinson, Albertslund, Denmark). PBMCs were thawed and washed before incubation with Fixable viability stain 620 (Becton Dickinson) and a set of conjugated monoclonal antibodies for 30 minutes on ice: CD3-APC-A750 (Beckmann Coulter), CD4-PE-Cy7 (Beckmann Coulter), CD8-A700 (Becton Dickinson), CD25-PE (Becton Dickinson), CD39-PerCP-Cy5.5 (Beckmann Coulter), CD196-BV510 (Biolegend, San Diego, USA), CD127-BV711 (Biolegend), CD161-BV650 (Becton Dickinson) and CD45RA-BV786 (Becton Dickinson). Next, PBMCs were washed and incubated with transcription factor buffer set (BD) for 45 min on ice. Afterwards, PBMCs were washed twice and intracellular monoclonal antibodies were added and incubated for 45 minutes on ice: RORγT-A488 (Becton Dickinson), FOXp3-A647 (Becton Dickinson) and Helios-PB (Beckmann Coulter). TH17 cells were determined by the CD4+RORγT+ phenotype, and T_reg_ cells by the CD4+CD25^high^FOXp3+ phenotype. Absolute cell counts in x10^9^/L were obtained by multiplying the frequency of T_H_17 and T_reg_ cells with the CD4+ T cell count from the same time point.

### Quantification of inflammation and infection markers

Markers were measured at the Department of Clinical Biochemistry, Copenhagen University Hospital Rigshospitalet, Denmark. As a marker of infection, plasma procalcitonin was determined by sandwich electrochemiluminescence immunoassays (ECLIA). As a marker of systemic inflammation, CRP was measured by latex immunoturbidimetric assays (LIA).

### DNA isolation from fecal, oral, and nasal samples and 16S rRNA gene sequencing

A total of 212 fecal samples for analysis of the intestinal microbiota were collected from 29 patients at the 10 time points described above. The gut microbiota was characterized at ≤6 time points in 9 patients (31%), at 7-8 time points in 13 patients (45%) and at 9-10 time points in 7 patients (24%) (Additional File 1: Table S1). DNA from fecal samples, one blank control per extraction round (thereof sequenced: 14), one mock community sample (Biodefense and Emerging Infectious Research (BEI) Resources of the American Type Culture Collection (ATCC) (Manassas, VA, USA), Catalog No. HM-276D) per sequencing run and two collection tube controls was isolated using the QIAamp Fast DNA Stool Mini kit (Qiagen, Venlo, Netherlands), following the manufacturer’s instructions with modifications according to [38].

We collected 248 buccal swabs (3x at ≤6 time points (10%), 11x at 7-8 time points (38%), 15x at 9-10 time points (52%)) and 249 anterior naris swabs (3x at ≤6 time points (10%), 9x at 7-8 time points (31%), 17x at 9-10 time points (59%)). DNA from swab samples, one blank control per extraction round (therof sequenced: 28), one mock community sample per run, two collection tube controls, and two sampling swab controls was isolated using the QIAamp UCP Pathogen Mini kit (Qiagen, Venlo, Netherlands), with the ‘Protocol: Pretreatment of Microbial DNA from Eye, Nasal, Pharyngeal, or other Swabs (Protocol without Pre-lysis)’ and subsequently the ‘Protocol: Sample Prep (Spin Protocol)’, following the manufacturer’s instructions with the following modifications: 550µl instead of 500µl Buffer ATL was used during pretreatment; DNA was eluted twice with 20µl Buffer AVE into 1.5 ml DNA LoBind tubes (Eppendorf, Hamburg, Germany) instead of the tubes provided with the kits.

Library construction and sequencing on an Illumina MiSeq instrument (Illumina Inc., San Diego, CA, USA) was performed at the Multi Assay Core facility (DMAC), Technical University of Denmark. DNA concentration of each sample was measured using a NanoDrop spectrophotometer (Thermo Scientific, Waltham, MA, USA). Library construction was performed according to the *16S Metagenomic Sequencing Library Preparation* protocol by Illumuna [39]: The V3-V4 region of the 16S ribosomal RNA gene were amplified in a PCR in each sample and in the controls, using the following previously evaluated primers, preceded by Illumina adapters [40]: 341F (5’-TCGTCGGCAGCGTCAGATGTGTATAAGAGACAGCCTACGGGNGGCWGCAG-3’) and 805R (5’-GTCTCGTGGGCTCGGAGATGTGTATAAGAGACAGGACTACHVGGGTATCTAATCC-3’). Amplicons were then analyzed for quantity and quality in an Agilent 2100 Bioanalyzer with the use of an Agilent RNA 1000 Nano Kit (Agilent Technology, Santa Clara, CA, USA). Subsequently, the amplicons were purified on AMPure XP Beads (Beckman Culter, Copenhagen, Denmark) according to the manufacturer’s instructions. Illumina adapters and dual-index barcodes were then added to the amplicon target in a PCR according to Illumina [39] using the 96 sample Nextera XT Index Kit (Illumina, FC-131–1002). A final clean-up of the libraries was performed in another PCR step, using AMPure XP Beads (Beckman Culter, Copenhagen, Denmark) according to the manufacturer’s instructions, followed by a confirmation of the target size in an Agilent 2100 Bioanalyzer (Agilent Technologies). Before sequencing, DNA concentration was determined with a Qubit (Life Technologies, Carlsbad, CA, USA) and libraries were pooled. In preparation for sequencing, the pooled libraries were denatured with NaOH, diluted with hybridization buffer, and heat denatured. 5% PhiX was included as an internal control for low-diversity libraries. Paired-end sequencing with 2 × 300bp reads was performed with a MiSeq v3 reagent kit on an Illumina MiSeq instrument (Illumina Inc., San Diego, CA, USA).

### 16S rRNA gene sequence pre-processing

Raw sequence reads were demultiplexed based on sample-specific barcodes and ‘read 1’ and ‘read 2’ FASTQ files for each sample were generated on the Illumina MiSeq instrument by the MiSeq reporter software. Primers were removed by using cutadapt (version 1.16) [41] at a tolerated maximum error rate of 15% for matching the primer sequence anchored in the beginning of each read. In the case that at least one read of a pair did not contain the primer, the pair was discarded. Only pairs in which the forward read contained the forward primer (341F) and the reverse read contained the reverse primer (805R) were retained.

The resulting reads were further processed using the R package DADA2 (version 1.8) to infer high-resolution amplicon sequence variants (ASV) [42]. Forward and reverse reads were truncated at 280 bp and 200 bp respectively. This way, the majority of reads retained a quality score >25 according to MultiQC analysis [43]. These truncation thresholds also ensured an overlap of 480 bp (expected amplicon length of 460 bp + 20 bp), allowing to merge forward and reverse reads. Samples were pooled for the sample inference step (*dada*() function) to increase the power for detecting rare variants. Default values were used for all other quality filtration parameters in DADA2. DNA from samples with a read count <10,000 after preliminary chimera and contaminant removal were re-sequenced. DNA from feces samples with a read count <5,000 were re-extracted. Eventually, chimeras were identified by sample and removed from the whole data set (over all sequencing runs) based on a consensus decision (*removeBimeraDenovo*() function, method “consensus”). Taxonomic assignment on ASVs was done by using the Silva reference data base (version 132), formatted for DADA2 [44]. Additional species assignment by exact reference strain matching was performed using the Silva species-assignment training data base, formatted for DADA2 [44].

The resulting ASV and taxonomy tables were integrated with the R package phyloseq and its dependencies (version 1.24.0) [45]. The data was split into two data sets, one containing feces sample data and one containing nasal and oral swab data. Subsequently, contaminant removal was performed with the R package decontam [46]. Potential technical batch effects by sequencing run, 96-well plate, extraction kit, extraction round, experimenter, and extraction date were assessed by ordination (Principal Coordinates Analysis (PCoA)).

For both, the fecal sample data set and the swab data set, contaminants were identified by sequencing run as a batch effect and a subsequent calculation of a consensus probability. For the feces sample data set, contaminants were identified by both, increased prevalence in 14 blank extraction controls and by relating ASV frequency to post-PCR sample DNA concentration, assuming inverse correlation (method “both”, frequency threshold: 0.2, prevalence threshold: 0.075) [46]. After manual evaluation of edge cases, 89 ASVs were removed from the fecal sample data set as contaminants. In an additional step, we identified 7 contaminants from 2 sampling tube controls (method and thresholds as stated above). In total, 96 ASVs were removed as contaminants from the fecal sample data set.

For the swab sample data set, contaminants were identified by both, increased prevalence in 28 blank extraction controls and by relating ASV frequency to post-PCR sample DNA concentration (method “both”, frequency threshold: 0.1, prevalence threshold: 0.6) [46]. A more stringent threshold for prevalence compared to frequency was chosen here, given the low biomass of the swab samples, accompanied by post-PCR DNA concentrations similar to those in blank controls. After manual evaluation of edge cases, 1137 ASVs were removed from the swab sample data set as contaminants. In an additional step, we identified 16 contaminants from 2 sampling tube controls and 2 swab controls (method “both”, frequency threshold: 0.075, prevalence threshold: 0.5). In total, 1153 ASVs were removed as contaminants from the oral and nasal swab sample data set.

For each subset, we created a phylogenetic tree by de novo alignment of the inferred ASVs, following a previously described workflow [47]. First, we performed multiple alignment with the package DECIPHER [48]. Subsequently, we built a neighbor-joining tree using the package phangorn [49], based on which we fitted a GTR+G+I (Generalized time-reversible with Gamma rate variation) maximum likelihood tree. The phylogenetic tree for each data set (fecal, oral, and nasal) was then integrated with the respective phyloseq object.

Next, we took core subsets of the ASVs remaining after contaminant removal using the function *kOverA*() from R package genefilter [50]. In the fecal set, 2465 ASVs with ≥5 reads in ≥2 samples were retained. With ≥5 reads in ≥10 samples, 509 ASVs were retained from the oral sample set, and 602 ASVs from the nasal sample set. Additional manual contaminant filtering was applied to the oral and nasal core sets. ASVs affiliated with taxonomic families commonly found in both the oral or nasal cavity and the gut were only retained in the oral sample set in case they had ≥10 reads in ≥10 samples. ASVs of families only expected in the gut were removed from the oral and nasal sample sets after manually assessing their abundances. Subsequently, we retained 377 ASVs in the oral sample set, and 197 ASVs in the nasal sample set.

For the comparison of the fecal microbiota in preexaminantion samples (n=15) of HSCT patients and healthy children (n=18), these data were combined in a phyloseq object. The same set of putative contaminants was removed from the healthy data set as were identified within the full fecal data set of HSCT patients. Subsequently, a core subset was taken as described above (retaining ASVs with ≥5 reads in ≥2 samples).

### Statistical analysis

Statistical analyses and generation of graphs was performed in R (version 3.5.1, R Foundation for Statistical Computing, Vienna, Austria) [51]. The R scripts documenting the major steps of our statistical analyses are available from figshare (https://doi.org/10.6084/m9.figshare.12280001). Sequencing data, and experimental and clinical data (https://doi.org/10.6084/m9.figshare.12280028) were integrated for analysis by using the R package phyloseq and its dependencies [45]. We also provide the resulting phyloseq objects through figshare (https://doi.org/10.6084/m9.figshare.12280004). Plots were generated with the packages ggplot2 [52], mixOmics [53], treeDA [54], caret [55], and partykit [56, 57]. From the core sets of ASV counts for each body site, bacterial alpha diversity (denoted by the inverse Simpson index) was calculated and compared between time points by using a Friedman test with Benjamini-Hochberg correction for multiple testing, and a post-hoc Conover test. To gain insight into changes of microbial abundances over time in relation to HSCT, we agglomerated ASV counts on family levels with the function *tax_glom()* in phyloseq [45]. Thereafter, we displayed the relative abundances of the 12 most abundant families at each body site for each time point. We also depicted relative abundances over time on family level in patients with aGvHD grade 0-I versus grade II-IV.

In order to determine which particular ASVs are relevant in temporal microbial abundance dynamics at each body site, we implemented tree-based sparse linear discriminant analysis (LDA) with the package treeDA [54]. This supervised method implements prior information about phylogenetic relationships between ASVs to perform supervised discrimination of classes, here time points, and induces sparsity constraints to increase interpretability [58]. Leaves and nodes of the phylogenetic tree, representing log+1 transformed ASV abundances and the sums thereof respectively, were used as predictive features. The core oral and nasal sets were used as input as described above, while the fecal set was further reduced to 389 ASVs with >5 reads in >10 samples for this analysis. Leave-one-out cross validation (LOOCV) was performed to choose the optimal minimum number of predictive features ensuring sparse, interpretable models. The resulting LDA models had 9 components. By default, this number corresponds to the number of predicted classes (here 10 time points) less one. To identify relevant components, we plotted sample scores colored by time points along each component and plotted the components pairwise against each other (Figure 1C). Thereby, we revealed that the first LDA-component for each body site showed the highest sample scores and best separated the samples by time point. Therefore, we proceeded with displaying temporal trajectories of clades of predictive features (ASVs) on the first component. For selected groups of predictive ASVs we displayed trajectories for patients with aGvHD grade 0-I versus with grade II-IV.

Next, we implemented machine learning models to predict aGvHD grade post-transplant from preceeding ASV abundances. The strategy and R code for the machine learning approach was partially adapted from a previous approach [59, 60]. As a preparative step for this analysis, we variance-stabilized the ASV count data. To do so, we first performed size factor estimation for zero-inflated data on the core data sets for each bbody site with the package GMPR [61]. Subsequently, we transformed the data by using the function *varianceStabilizingTransformation*() in the package DESeq2 [62]. The function implements a Gamma-Poisson mixture model to account for both library size differences and biological variability [63]. For the prediction of aGvHD grade, we compared the performances of four different classifiers (random forest (rf), boosted logistic regression (LogitBoost), support vector machines with linear kernel (svmLinear), and support vector machines with radial basis function kernel (svmRadial)) using the package caret [55]. We took subsets of the phyloseq objects comprising only the time points preceding aGvHD onset: pre-examination, conditioning start, and at the time of HSCT. Prior to fitting the models, we excluded ASVs with near zero variance, i.e. those that were not differentially abundant between any samples, by using the function *nearZeroVar*() in package caret [55]. Thereby we obtained sets of 238, 186, and 100 ASVs for the fecal, oral, and nasal data set, respectively, which were then assessed as potential predictors of subsequent aGvHD. All classifiers were trained on a randomly chosen subset of 70% of the data to build a predictive model evaluated on a test set (30% of the data). Splitting was performed in a way that samples from the same patient at different time points were kept together in either the testing or training set to ensure that the outcome of a patient can only appear in either the testing set or the training set, but not both. Thirty iterations of 10-fold cross-validation were performed for each classifier, both with and without up-sampling. Up-sampling refers to the process of replacement-based sampling of the class with fewer samples (here aGvHD grade II-IV) to the same size as the class with more samples (here aGvHD grade 0-I) to achieve a balanced design. SvmLinear on up-sampled data was chosen as the best performing predictive model for all three data sets (gut, oral, and nasal). Subsequently, we performed Boruta feature selection using the package Boruta [64]. The Boruta algorithm is a Random Forest classification based wrapper that compares the importance of real features to that of so called ‘shadow attributes’ with randomly shuffled values. Features that are less important than the ‘shadow attributes’ are iteratively removed. Here, we retained those ASVs in each data set that were both, among the 50 most important predictors in the svmLinear model and confirmed by the Boruta algorithm (Additional File 1: Table S3). Subsequently, we fitted a CTREE on each set of selected predictors (17 gut, 26 oral, and 12 nasal ASVs) by using the package partykit (Additional File 1: Table S3) [56, 57]. In the CTREE analysis the effect of the predictive ASVs on aGvHD grade is evaluated in a nonparametric regression framework. Using CTREE, we found 3 significant ASVs each in the gut and in the oral data set, and two significant ASVs in the nasal data set. CTREE iteratively tests if the abundance of any ASV has a significant effect on aGvHD grade.

In the case that a significant relation is found, the ASVs with the largest effect is picked as a node for the tree. The procedure is then recursively repeated until no further significant effect of any ASV on aGvHD is found. We plotted the result as a tree featuring the significant split nodes, represented by the ASVs and the Bonferroni-corrected p-values indication significant influence of their abundance on aGvHD grade. The terminal nodes of the tree show the proportion of samples stemming from patients with aGvHD grade 0-I versus II-IV, under the condition of the abundance split criterion described on each branch. Since we used variance stabilized bacterial abundances as input for the machine learning analyses, abundances can be presented as negative values in some cases and are therefore not easy to interpret intuitively. Therefore, we additionally displayed the log-transformed relative abundances of all ASVs significantly predicting aGvHD in boxplots at the three investigated time points (pre-examination, conditioning start, and at the time of HSCT).

Subsequently, we were interested in associations between the fecal, oral, and nasal microbiota and immune cell counts, and clinical outcomes in HSCT. Records of immune markers, and immune cell counts contained left- and right-censored measurements, i.e. observations below or above the detection (or recording) limit, respectively. In order to use these data in analyses that do not tolerate censored records, we needed to impute the censored data. Therefore, we first fitted the non-parametric maximum likelihood estimator (NPMLE, also called Turnbull estimator) for univariate interval censored data on each variable that contained censored records, using the function *ic_np*() in the R package icenReg [65]. Subsequently, censored records were imputed, informed by the model that was fitted on the entity of observed and censored data of each variable, using the *imputeCens()* function [65]. Next, we took the median of measurements for the time points defined above for those immune markers, and immune cell counts that have been measured more frequently than that. This way, we obtained comparable data sets. Continuous immune marker and cell count data that was systematically missing for certain sampling time points was split by time points and unavailable time points were excluded. Missing values in continuous immune marker and cell count data were imputed for variables with ≤ 50% missingness. Simultaneous multivariate non-parametric imputation was performed using the R package missForest [66]. Variables with more than 50% missing values were excluded from the analysis.

Next, we implemented two multivariate multi-table approaches to gain a detailed understanding of how the fecal, oral, and nasal microbiota might be associated with immune cell counts, immune markers, and clinical outcomes in HSCT. Evaluated clinical outcomes comprised acute GvHD (grade 0-I versus II-IV), relapse, overall survival, and treatment-related mortality. Furthermore, we included bacterial alpha diversity (inversed Simpson index), antibiotic treatment, infections, Karnofsky scores before conditioning and at day +100, and patients’ baseline parameters (age, weight, sex, primary disease, malignant versus benign primary disease, conditioning regimen (including ATG treatment), chemotherapeutic agents’ dosages, TBI treatment and dosage, stem cell source, GvHD prophylactic regimen, donor type (sibling/matched unrelated/haploidentical), donor HLA-match, and donor sex).

For each body site, we performed sparse partial least squares (sPLS) regression by using the function *spls*() in the package mixOmics [53]. In sPLS regression, two matrices are being integrated and both their structures are being modelled. Here, we used variance stabilized ASV abundances as explanatory variables and all continuous clinical and immune parameters as response variables. The method allows multiple response variables. Collinear, and noisy data can be handled by this method as well [67]. We did not limit the number of response variables to be kept for each component (keepY) prior to model calculation. The number of explanatory variables (ASVs) to be kept on each component (keepX) was set to 25 after running the sPLS regression models for each body site with a range of values between 20 and 40 for keepX, showing results robust to keepX. The *perf()* function was used to inform the choice of 3 relevant components. Based on the sPLS regression models for each body site, we then performed hierarchical clustering with the *cim()* function, using the clustering method “complete linkage” and the distance method “Pearson’s correlation”. Thereby, we generated matrices of coefficients indicating correlations between ASV abundances and continuous clinical and immune parameters.

Subsequently, we carried out canonical (i.e. bidirectional) correspondence analysis (CCpnA), which is a multivariate constrained ordination method. This method allow us to assess associations of both categorical and continuous clinical and immune parameters to ASV abundances. We included ASVs and variables with a correlation of >0.2/<-0.2 (oral and nasal data set) or >0.3/<-0.3 (fecal data set) in the sPLS analysis into the CCpnA, and additionally included categorical variables that could not be included in the sPLS. The method was implemented with the *cca()* function in package vegan [68]. It implements a Chi-square transformation of the log+1 transformed ASV count matrix and subsequent weighted linear regression, followed by singular value decomposition. We depicted the CCpnA results as a triplot with plot dimensions corresponding in length to the percentage of variance explained by each axis. At each body site, we identified three clusters of ASVs through hierarchical clustering based on the first three latent dimensions of each sPLS analysis (Figure 6A, and Additional File 2: Figures S5A and S6A). The CCpnA analyses reinforced the cluster separations and additionally provided insight into associations with categorical variables, including patient baseline parameters, the occurrence of infections, antibiotics treatment, and clinical outcomes (Figure 6B, and Additional File 2: Figures S5B and S6B).

We compared bacterial alpha diversity and community composition in the gut of HSCT patients at preexamination with that of healthy children. Alpha diversity (inverse Simpson index) between the two groups was compared by a Kruskal-Wallis test. Community composition was visualized in a principal coordinates analysis (PCoA), and analysis of similarities (ANOSIM, package vegan) was used to assess significant differences in the means of rank dissimilarities between the two groups. DESeq2 was employed for identification of differentially abundant genera among the top 100 most abundant genera with >10 total reads [62]. Differences in relative abundance of genera identified as differentially abundant were visualized in a heat tree (package metacoder) [69]. Higher taxonomic level differential abundance was assessed by linear discriminant analysis effect size (LEfSe) on centered-log ratio (CLR) transformed data with an LDA cutoff of 4 (package microbiomeMarker) [70]. LefSe accounts for the hierarchical structure of bacterial phylogeny, thereby allowing identification of differentially abundant taxa on several taxonomic levels (here kingdom to genus). For additional information see https://doi.org/10.6084/m9.figshare.13614230).

## Data Availability

The 16S rRNA gene sequences are available through the European Nucleotide Archive (ENA) at the European Bioinformatics Institute (EBI) under accession number PRJEB30894. The datasets generated and/or analysed in this study as well as the R code used to analyze the data are available from the figshare repository https://figshare.com/projects/Microbiota_long-term_dynamics_and_prediction_of_acute_graft-versus-host-disease_in_allogeneic_stem_cell_transplantation/80366 (see also individual links in the Methods section).

## List of abbreviations

AML: Acute myeloid leukemia
ASV: Amplicon sequence variant
ATG: Anti-thymocyte globulin
CCpnA: Canonical correspondence analysis
CML: Chronic myeloid leukemia
CRP: C-reactive protein
CTREE: Conditional inference tree
ECLIA: Electrochemiluminescence immunoassays
GvL effect: Graft-versus-leukemia effect
(a)GvHD: (Acute) graft-versus-host disease
HSCT: Hematopoietic stem cell transplantation
IDS: Immunodeficiency syndromes
IEA: Inherited abnormalities of erythrocyte differentiation or function
IMD: Inherited disorders of metabolism
LIA: Latex immunoturbidimetric assay
LDA: Linear discriminant analysis
LogitBoost: Boosted logistic regression
LOOCV: Leave-one-out cross validation
MDS: Myelodysplastic or myeloproliferative disorders
MM: Multiple myeloma
NHL: Non-Hodgkin lymphomas
NPMLE: Non-parametric maximum likelihood estimator
OL: Other leukemia
OTU: Operational taxonomic unit
PBMC: Peripheral blood mononuclear cell
PCoA: Principal Coordinates Analysis
Rf: Random forest
SAA: Severe aplastic anemia
SCFA: Short-chain fatty acid
sPLS: Sparse partial least squares analysis
svmLinear: Support vector machines with linear kernel
svmRadial: Support vector machines with radial basis function kernel
TBI: Total body irradiation
T_H_17 cell: T helper 17 cell
T_reg_ cell: T regulatory cell
UCB: Umbilical cord blood

## Acknowledgements

We thank the patients and their families for their participation in this study. We also thank Marlene Danner Dalgaard and Neslihan Bicen (Multi Assay Core facility (DMAC), Technical University of Denmark) for library construction and sequencing. Sequence pre-processing described in this paper was performed using the DeiC National Life Science Supercomputer at DTU. Furthermore, we would like to thank Patrick Murigu Kamau Njage (Technical University of Denmark) for helpful discussions related to machine learning models.

## Declarations

### Author’s contributions

A.C.I., K.K., K.G.M., and S.J.P. designed the research; A.C.I., K.K., H.M, M.I., and S.J.P. performed the research; A.C.I., and S.J.P. contributed analytic tools; A.C.I., and S.J.P. analyzed the data; A.C.I. and S.J.P. wrote the manuscript; and K.K., M.I., F.M.A., and K.G.M. edited the manuscript.

### Ethics approval and consent to participate

Written informed consent was obtained from the patients and/or their legal guardians. The study protocol was approved by the local ethics committee (H-7-2014-016) and the Danish Data Protection Agency.

### Consent for publication

Not applicable.

---

**Supplementary Table S1. Patient characteristics.** Abbreviations: HLA, human leukocyte antigen; TBI, total body irradiation; CY, Cyclophosphamide; VP16, Etoposide; BU, Busulfan; MEL, Melphalan; GvHD, graft-versus-host disease.

**Supplementary Table S2. Taxonomy of a subset of LDA clade members and corresponding LDA-coefficients in the gut, oral cavity, and nasal cavity.**

**Supplementary Table S3. Taxonomy of aGvHD predictors within the fecal, oral, and nasal microbiota.** ASVs that were significantly predicting aGvHD severity according to the conditional inference tree regression model are highlighted in bold. Of the 50 most important gut ASVs identified by the svmLinear model, 17 were confirmed by Boruta feature selection and are listed here. In the oral and nasal cavities, 26 and 12 ASVs were confirmed by Boruta selection, respectively. Listed in bold are those ASVs with a significant predictive effect on aGvHD severity, tested in a regression framework with CTREE (see Methods).

**Figure S1. The gut microbiota in the HSCT patients at pre-exam differs from the gut microbiota of age-matched healthy children.** A) Fecal bacterial alpha diversity (inverse Simpson index) was 2.4-fold higher in healthy children (n=18) compared to children at pre-examination before HSCT (n=15). B) Fecal bacterial composition was significantly different between the two groups (anosim, p=0.001, R=0.44), and within-group variance was significantly greater in the HSCT group (betadisper, p<0.001). C) The taxa which best explain differences in community structure between HSCT patients at preexamination and healthy children were identified by analysis of LEfSe (Linear discriminant analysis Effect Size). LefSe accounts for the hierarchical structure of bacterial phylogeny, thereby allowing identification of differentially abundant taxa on several taxonomic levels (here: kingdom to genus). Count data was centered-log ratio (CLR) transformed within the LEfSe analysis. The higher the LDA score (log10), the higher the effect size of the respective taxon in explaining group difference. Here, we show taxa with an LDA score >4. D) Differentially abundant genera between the two groups were additionally identified by DESeq2. Of the top 100 most abundant genera (of the whole gut microbiota data set), eighteen genera were significantly more abundant in healthy children (yellow), and 15 genera were significantly more abundant in the patients at preexam (purple). Differences in median proportions of these genera (and their supertaxa) are displayed in a heat tree. See also additional information at https://doi.org/10.6084/m9.figshare.13614230.

**Figure S2. Most abundant taxonomic families in the gut, oral cavity, and nasal cavity in allo-HSCT patients.** Rank abundance curves displaying the proportions of the 12 most abundant taxonomic families at each body site (gut, oral cavity, and nasal cavity).

**Figure S3. Tree-based sparse linear discriminant analysis revealing nasal ASVs that distinguish time points from each other in relation to HSCT.** A) Relative abundances over time of the 12 most abundant families in the nasal cavity. B) Coefficients of discriminating clades of ASVs on the first LDA axis, colored by taxonomic family, and plotted along the phylogenetic tree. C) Trajectories of ASVs in one discriminating group, affiliated with the family *Corynebacteriaceae,* with decreasing abundances after HSCT and recovery at late follow-up time points. The most abundant discriminating ASV is indicated Detailed taxonomic information and LDA-coefficients of the displayed ASVs are listed in Table S2.

**Figure S4. Machine learning-based prediction of aGvHD severity from nasal microbial abundances pre-HSCT.** A) Relative abundances of the 12 most abundant families over time in the nasal cavity in patients with aGvHD grade 0-I versus II-IV. B) Importance plot of top 20 predictive nasal ASVs identified by the svmLinear model with importance scores indicating the mean decrease in prediction accuracy in case the respective ASV would be excluded from the model. The final cross-validated svmLinear model predicted aGvHD (0-I versus II-IV) from the abundances of nasal ASVs pre-HSCT with 76% accuracy (95% CI: 56% to 90%). The ASVs that were also confirmed by Boruta feature selection are indicated with asterisk. C) Conditional inference tree (CTREE) displaying ASVs identified as significant split nodes by nonparametric regression for prediction of aGvHD. Numbers along the branches indicate split values of variance stabilized bacterial abundances. The terminal nodes show the proportion of samples originating from patients with aGvHD grade 0-I vs II-IV (n = number of samples). D) Boxplots depict the log transformed relative abundances of the predictive ASVs at time points up to the transplantation in aGvHD grade 0-I compared with grade II-IV patients.

**Figure S5. Multivariate associations of the oral microbiota with immune and clinical parameters in HSCT.** A) Clustered image map (CIM) based on sparse partial least squares (sPLS) regression analysis dimensions 1, 2, and 3, displaying pairwise correlations >0.2/<-0.2 between oral ASVs (bottom), and continuous immune and clinical parameters (right). Red indicated positive correlation, and blue indicates negative correlation, respectively. Based on the sPLS regression model, hierarchical clustering (clustering method: complete linkage, distance method: Pearson’s correlation) was performed resulting in the three depicted clusters. B) Canonical correspondence analysis (CCpnA) relating oral microbial abundances (circles) to continuous (arrows) and categorical (+) immune and clinical parameters. ASVs and variables with at least one correlation >0.2/<-0.2 in the sPLS analysis were included in the CCpnA. The triplot shows variables and ASVs with a score >0.3/<-0.3 on at least one the first three CCpnA axes, displayed on axis 1 versus 2 with samples depicted as triangles. The colored ellipses (depicted with 80% confidence interval) correspond to the clusters of ASVs identified by the sPLS-based hierarchical clustering. For visualization purposes, a focused section of the CCpnA triplot is shown. Abbreviations are described in Figure 6. Additional abbreviations: fungal, fungal infection; haploident, haploidentical donor; hemo, hemoglobin; leuko, leukocytes; lympho, lymphocytes; w1, week+1; w2, week+2; w3, week+3.

**Figure S6. Multivariate associations of the nasal microbiota with immune and clinical parameters in HSCT.** A) Clustered image map (CIM) based on sparse partial least squares (sPLS) regression analysis dimensions 1, 2, and 3, displaying pairwise correlations >0.2/<-0.2 between nasal ASVs (bottom), and continuous immune and clinical parameters (right). Red indicated positive correlation, and blue indicates negative correlation, respectively. Based on the sPLS regression model, hierarchical clustering (clustering method: complete linkage, distance method: Pearson’s correlation) was performed resulting in the three depicted clusters. B) Canonical correspondence analysis (CCpnA) relating nasal microbial abundances (circles) to continuous (arrows) and categorical (+) immune and clinical parameters. ASVs and variables with at least one correlation >0.2/<-0.2 in the sPLS analysis were included in the CCpnA. The triplot shows variables and ASVs with a score >0.3/<-0.3 on at least one the first three CCpnA axes, displayed on axis 1 versus 2 with samples depicted as triangles. The colored ellipses (depicted with 80% confidence interval) correspond to the clusters of ASVs identified by the sPLS-based hierarchical clustering. For visualization purposes, a focused section of the CCpnA triplot is shown. Abbreviations are described in Figures 6 and S5. Additional abbreviations: DonorMatch8, unrelated donor with 1 HLA mismatch; PB, peripheral blood.

## References

1. Chabannon C, Kuball J, Bondanza A, Dazzi F, Pedrazzoli P, Toubert A, et al. Hematopoietic stem cell transplantation in its 60s: A platform for cellular therapies. Sci Transl Med [Internet]. American Association for the Advancement of Science; 2018 [cited 2018 Aug 2];10:eaap9630. Available from: http://www.ncbi.nlm.nih.gov/pubmed/29643233

2. Shono Y, van den Brink MRM. Gut microbiota injury in allogeneic haematopoietic stem cell transplantation. Nat Rev Cancer [Internet]. Nature Publishing Group; 2018 [cited 2018 Feb 21]; Available from: http://www.nature.com/doifinder/10.1038/nrc.2018.10

3. Holler E, Butzhammer P, Schmid K, Hundsrucker C, Koestler J, Peter K, et al. Metagenomic Analysis of the Stool Microbiome in Patients Receiving Allogeneic Stem Cell Transplantation: Loss of Diversity Is Associated with Use of Systemic Antibiotics and More Pronounced in Gastrointestinal Graft-versus-Host Disease. Biol Blood Marrow Transplant [Internet]. 2014 [cited 2015 Oct 22];20:640–5. Available from: http://www.sciencedirect.com/science/article/pii/S1083879114000755

4. Ingham AC, Kielsen K, Cilieborg MS, Lund O, Holmes S, Aarestrup FM, et al. Specific gut microbiome members are associated with distinct immune markers in pediatric allogeneic hematopoietic stem cell transplantation. Microbiome [Internet]. BioMed Central; 2019 [cited 2019 Sep 18];7:131. Available from: https://microbiomejournal.biomedcentral.com/articles/10.1186/s40168-019-0745-z

5. Rivera-Chávez F, Lopez CA, Bäumler AJ. Oxygen as a driver of gut dysbiosis. Free Radic Biol Med [Internet]. Pergamon; 2017 [cited 2018 Feb 18];105:93–101. Available from: https://www.sciencedirect.com/science/article/pii/S0891584916304361?via%3Dihub

6. Taur Y, Xavier JB, Lipuma L, Ubeda C, Goldberg J, Gobourne a., et al. Intestinal Domination and the Risk of Bacteremia in Patients Undergoing Allogeneic Hematopoietic Stem Cell Transplantation. Clin Infect Dis [Internet]. 2012;55:905–14. Available from: http://cid.oxfordjournals.org/lookup/doi/10.1093/cid/cis580

7. Ghimire S, Weber D, Mavin E, Wang X nong, Dickinson AM, Holler E. Pathophysiology of GvHD and Other HSCT-Related Major Complications. Front Immunol [Internet]. Frontiers; 2017 [cited 2018 Oct 19];8:79. Available from: http://journal.frontiersin.org/article/10.3389/fimmu.2017.00079/full

8. Golob JL, Pergam SA, Srinivasan S, Fiedler TL, Liu C, Garcia K, et al. Stool Microbiota at Neutrophil Recovery Is Predictive for Severe Acute Graft vs Host Disease After Hematopoietic Cell Transplantation. Clin Infect Dis [Internet]. Oxford University Press; 2017 [cited 2018 Nov 23];65:1984–91. Available from: https://academic.oup.com/cid/article/65/12/1984/4085173

9. Jenq RR, Taur Y, Devlin SM, Ponce DM, Goldberg JD, Ahr KF, et al. Intestinal Blautia Is Associated with Reduced Death from Graft-versus-Host Disease. Biol Blood Marrow Transplant [Internet]. 2015 [cited 2016 May 9];21:1373–83. Available from: http://www.sciencedirect.com/science/article/pii/S1083879115002931

10. Han L, Zhao K, Li Y, Han H, Zhou L, Ma P, et al. A gut microbiota score predicting acute graft-versus-host disease following myeloablative allogeneic hematopoietic stem cell transplantation. Am J Transplant. 2020;20:1014–27.

11. Peled JU, Gomes ALC, Devlin SM, Littmann ER, Taur Y, Sung AD, et al. Microbiota as predictor of mortality in allogeneic hematopoietic-cell transplantation. N Engl J Med. 2020;382:822–34.

12. Stein-Thoeringer CK, Nichols KB, Lazrak A, Docampo MD, Slingerland AE, Slingerland JB, et al. Lactose drives Enterococcus expansion to promote graft-versus-host disease. Science (80-). 2019;366:1143–9.

13. Honda K, Littman DR. The microbiota in adaptive immune homeostasis and disease. Nature [Internet]. Nature Publishing Group; 2016 [cited 2018 Aug 21];535:75–84. Available from: http://www.nature.com/articles/nature18848

14. Atarashi K, Tanoue T, Oshima K, Suda W, Nagano Y, Nishikawa H, et al. Treg induction by a rationally selected mixture of Clostridia strains from the human microbiota. Nature [Internet]. Nature Publishing Group; 2013 [cited 2018 Sep 6];500:232–6. Available from: http://www.nature.com/articles/nature12331

15. Kielsen K, Ryder LP, Lennox-Hvenekilde D, Gad M, Nielsen CH, Heilmann C, et al. Reconstitution of Th17, Tc17 and Treg cells after paediatric haematopoietic stem cell transplantation: Impact of interleukin-7. Immunobiology [Internet]. 2018 [cited 2018 Feb 7];223:220–6. Available from: http://www.ncbi.nlm.nih.gov/pubmed/29033080

16. Han L, Jin H, Zhou L, Zhang X, Fan Z, Dai M, et al. Intestinal Microbiota at Engraftment Influence Acute Graft-Versus-Host Disease via the Treg/Th17 Balance in Allo-HSCT Recipients. Front Immunol [Internet]. 2018 [cited 2018 May 17];9:669. Available from: http://www.ncbi.nlm.nih.gov/pubmed/29740427

17. Ratajczak P, Janin A, Peffault de Latour R, Leboeuf C, Desveaux A, Keyvanfar K, et al. Th17/Treg ratio in human graft-versus-host disease. Blood [Internet]. American Society of Hematology; 2010 [cited 2018 Nov 19];116:1165–71. Available from: http://www.ncbi.nlm.nih.gov/pubmed/20484086

18. Larsen JM. The immune response to *Prevotella* bacteria in chronic inflammatory disease. Immunology [Internet]. Wiley/Blackwell (10.1111); 2017 [cited 2018 Nov 25];151:363–74. Available from: http://doi.wiley.com/10.1111/imm.12760

19. De Pietri S, Ingham AC, Frandsen TL, Rathe M, Krych L, Castro-Mejía JL, et al. Gastrointestinal toxicity during induction treatment for childhood acute lymphoblastic leukemia: The impact of the gut microbiota. Int J Cancer. Wiley-Liss Inc.; 2020;147:1953–62.

20. Weber D, Oefner PJ, Hiergeist A, Koestler J, Gessner A, Weber M, et al. Low urinary indoxyl sulfate levels early after transplantation reflect a disrupted microbiome and are associated with poor outcome. Blood [Internet]. 2015 [cited 2015 Oct 22];126:1723–8. Available from: http://www.bloodjournal.org/content/126/14/1723

21. Taur Y, Jenq RR, Perales M, Littmann ER, Morjaria S, Ling L, et al. The effects of intestinal tract bacterial diversity on mortality following allogeneic hematopoietic stem cell transplantation. Transplantation [Internet]. 2014;124:1174–82. Available from: http://www.bloodjournal.org/content/bloodjournal/124/7/1174.full.pdf?sso-checked=true

22. Andermann TM, Peled JU, Ho C, Reddy P, Riches M, Storb R, et al. The Microbiome and Hematopoietic Cell Transplantation: Past, Present, and Future. Biol Blood Marrow Transplant [Internet]. Elsevier; 2018 [cited 2018 May 29]; Available from: https://www.sciencedirect.com/science/article/pii/S1083879118300879?via%3Dihub

23. Jenq RR, Ubeda C, Taur Y, Menezes CC, Khanin R, Dudakov J a., et al. Regulation of intestinal inflammation by microbiota following allogeneic bone marrow transplantation. J Exp Med [Internet]. 2012;209:903–11. Available from: http://www.jem.org/cgi/doi/10.1084/jem.20112408

24. Verma D, Garg PK, Dubey AK. Insights into the human oral microbiome. Arch Microbiol [Internet]. Springer Berlin Heidelberg; 2018;200:525–40. Available from: http://dx.doi.org/10.1007/s00203-018-1505-3

25. Osakabe L, Utsumi A, Saito B, Okamatsu Y, Kinouchi H, Nakamaki T, et al. Influence of Oral Anaerobic Bacteria on Hematopoietic Stem Cell Transplantation Patients: Oral Mucositis and General Condition. Transplant Proc [Internet]. Elsevier; 2017 [cited 2018 Mar 12];49:2176–82. Available from: http://www.ncbi.nlm.nih.gov/pubmed/29149979

26. Soga Y, Maeda Y, Ishimaru F, Tanimoto M, Maeda H, Nishimura F, et al. Bacterial substitution of coagulase-negative staphylococci for streptococci on the oral mucosa after hematopoietic cell transplantation. Support Care Cancer. 2011;19:995–1000.

27. Olczak-Kowalczyk D, Daszkiewicz M, Krasuska-Slawińska, Dembowska-Bagińska B, Gozdowski D, Daszkiewicz P, et al. Bacteria and Candida yeasts in inflammations of the oral mucosa in children with secondary immunodeficiency. J Oral Pathol Med. 2012;41:568–76.

28. Chung H, Pamp SJ, Hill JA, Surana NK, Edelman SM, Troy EB, et al. Gut immune maturation depends on colonization with a host-specific microbiota. Cell [Internet]. Elsevier; 2012 [cited 2018 Aug 16];149:1578–93. Available from: http://www.ncbi.nlm.nih.gov/pubmed/22726443

29. Mathewson ND, Jenq R, Mathew A V, Koenigsknecht M, Hanash A, Toubai T, et al. Gut microbiome-derived metabolites modulate intestinal epithelial cell damage and mitigate graft-versus-host disease. Nat Immunol [Internet]. NIH Public Access; 2016 [cited 2018 May 15];17:505–13. Available from: http://www.ncbi.nlm.nih.gov/pubmed/26998764

30. Kim M, Qie Y, Park J, Kim CH. Gut Microbial Metabolites Fuel Host Antibody Responses. Cell Host Microbe [Internet]. Elsevier Inc.; 2016;20:202–14. Available from: http://dx.doi.org/10.1016/j.chom.2016.07.001

31. Shono Y, Docampo MD, Peled JU, Perobelli SM, Velardi E, Tsai JJ, et al. Increased GVHD-related mortality with broad-spectrum antibiotic use after allogeneic hematopoietic stem cell transplantation in human patients and mice. Sci Transl Med [Internet]. 2016 [cited 2016 May 23];8:339ra71–339ra71. Available from: http://stm.sciencemag.org/content/8/339/339ra71

32. Weber D, Jenq RR, Peled JU, Taur Y, Hiergeist A, Koestler J, et al. Microbiota Disruption Induced by Early Use of Broad Spectrum Antibiotics is an Independent Risk Factor of Outcome after Allogeneic Stem Cell Transplantation [Internet]. Biol. Blood Marrow Transplant. Elsevier Inc.; 2017. Available from: http://linkinghub.elsevier.com/retrieve/pii/S1083879117302756

33. Weber D, Hiergeist A, Weber M, Dettmer K, Wolff D, Hahn J, et al. Detrimental effect of broad-spectrum antibiotics on intestinal microbiome diversity in patients after allogeneic stem cell transplantation: Lack of commensal sparing antibiotics. Clin Infect Dis [Internet]. 2018 [cited 2018 Sep 20]; Available from: http://www.ncbi.nlm.nih.gov/pubmed/30124813

34. Liu C, Frank DN, Horch M, Chau S, Ir D, Horch EA, et al. Associations between acute gastrointestinal GvHD and the baseline gut microbiota of allogeneic hematopoietic stem cell transplant recipients and donors. Bone Marrow Transplant Adv online Publ [Internet]. 2017;doi:1–8. Available from: https://www.nature.com/bmt/journal/vaop/ncurrent/pdf/bmt2017200a.pdf

35. Biagi E, Zama D, Nastasi C, Consolandi C, Fiori J, Rampelli S, et al. Gut microbiota trajectory in pediatric patients undergoing hematopoietic SCT. Bone Marrow Transplant [Internet]. Nature Publishing Group; 2015 [cited 2018 Jul 2];50:992–8. Available from: http://www.nature.com/articles/bmt201516

36. Mancini N, Greco R, Pasciuta R, Barbanti MC, Pini G, Morrow OB, et al. Enteric Microbiome Markers as Early Predictors of Clinical Outcome in Allogeneic Hematopoietic Stem Cell Transplant: Results of a Prospective Study in Adult Patients. Open Forum Infect Dis [Internet]. Oxford University Press; 2017 [cited 2018 Dec 8];4. Available from: http://academic.oup.com/ofid/article/doi/10.1093/ofid/ofx215/4367678

37. Glucksberg H, Storb R, Fefer A, Buckner CD, Neiman PE, Clift RA, et al. Clinical manifestations of graft-versus-host disease in human recipients of marrow from HL-A-matched sibling donors. Transplantation [Internet]. 1974;18:295–304. Available from: http://www.ncbi.nlm.nih.gov/pubmed/4153799

38. Knudsen BE, Bergmark L, Munk P, Lukjancenko O, Prieme A, Aarestrup FM, et al. Impact of Sample Type and DNA Isolation Procedure on Genomic Inference of Microbiome Composition. bioRxiv [Internet]. 2016;1:064394. Available from: http://biorxiv.org/lookup/doi/10.1101/064394

39. 16S Metagenomic Sequencing Library Preparation. [Internet]. [cited 2018 Apr 17]. Available from: https://support.illumina.com/content/dam/illumina-support/documents/documentation/chemistry_documentation/16s/16s-metagenomic-library-prep-guide-15044223-b.pdf

40. Klindworth A, Pruesse E, Schweer T, Peplies J, Quast C, Horn M, et al. Evaluation of general 16S ribosomal RNA gene PCR primers for classical and next-generation sequencing-based diversity studies. Nucleic Acids Res [Internet]. Oxford University Press; 2013 [cited 2018 Apr 17];41:e1–e1. Available from: http://academic.oup.com/nar/article/41/1/e1/1164457/Evaluation-of-general-16S-ribosomal-RNA-gene-PCR

41. Martin M. Cutadapt removes adapter sequences from high-throughput sequencing reads. EMBnet.journal [Internet]. 2011 [cited 2018 Jun 26];17:10. Available from: http://journal.embnet.org/index.php/embnetjournal/article/view/200

42. Callahan BJ, McMurdie PJ, Rosen MJ, Han AW, Johnson AJA, Holmes SP. DADA2: High-resolution sample inference from Illumina amplicon data. Nat Methods [Internet]. 2016 [cited 2016 Jul 28];13:581–3. Available from: http://www.nature.com/nmeth/journal/v13/n7/full/nmeth.3869.html

43. Ewels P, Magnusson M, Lundin S, Käller M. MultiQC: summarize analysis results for multiple tools and samples in a single report. Bioinformatics [Internet]. Oxford University Press; 2016 [cited 2018 Sep 11];32:3047–8. Available from: https://academic.oup.com/bioinformatics/article-lookup/doi/10.1093/bioinformatics/btw354

44. Callahan B. Silva taxonomic training data formatted for DADA2 (Silva version 132). 2018 [cited 2018 Jun 26]; Available from: https://doi.org/10.5281/zenodo.1172783#.WzJRh15uQOA.mendeley

45. McMurdie PJ, Holmes S. phyloseq: An R Package for Reproducible Interactive Analysis and Graphics of Microbiome Census Data. Watson M, editor. PLoS One [Internet]. Public Library of Science; 2013 [cited 2018 Jan 24];8:e61217. Available from: http://dx.plos.org/10.1371/journal.pone.0061217

46. Davis NM, Proctor DM, Holmes SP, Relman DA, Callahan BJ. Simple statistical identification and removal of contaminant sequences in marker-gene and metagenomics data. Microbiome [Internet]. BioMed Central; 2018 [cited 2018 Dec 27];6:226. Available from: https://microbiomejournal.biomedcentral.com/articles/10.1186/s40168-018-0605-2

47. Callahan BJ, Sankaran K, Fukuyama JA, McMurdie PJ, Holmes SP. Bioconductor workflow for microbiome data analysis: from raw reads to community analyses. F1000Research [Internet]. 2016;5:1492. Available from: http://www.ncbi.nlm.nih.gov/pubmed/27508062 %5Cn http://www.pubmedcentral.nih.gov/articlerender.fcgi?artid=PMC4955027

48. Wright ES. Using DECIPHER v2.0 to Analyze Big Biological Sequence Data in R. R J. 2016;8:352–9.

49. Schliep KP. phangorn: phylogenetic analysis in R. Bioinformatics [Internet]. Oxford University Press; 2011 [cited 2018 Nov 27];27:592–3. Available from: http://www.ncbi.nlm.nih.gov/pubmed/21169378

50. Gentleman R, Carey V, Huber W, Hahne F. genefilter: methods for filtering genes from microarray experiments. R package version 1.58.1. 2017.

51. R Core Team. R: A Language and Environment for Statistical Computing [Internet]. R Foundation for Statistical Computing, Vienna, Austria; 2018. Available from: https://www.r-project.org/

52. . Wickham H. ggplot2: Elegant Graphics for Data Analysis. Springer Verlag New York; 2016.

53. Rohart F, Gautier B, Singh A, Lê Cao K-A. mixOmics: An R package for ‘omics feature selection and multiple data integration. Schneidman D, editor. PLOS Comput Biol [Internet]. Public Library of Science; 2017 [cited 2017 Dec 11];13:e1005752. Available from: http://dx.plos.org/10.1371/journal.pcbi.1005752

54. Fukuyama J. treeDA: Tree-Based Discriminant Analysis. [Internet]. 2017. Available from: https://github.com/jfukuyama/treeda

55. Kuhn M, Wing J, Weston S, Williams A, Keefer C, Engelhardt A, et al. caret: Classification and Regression Training. R package version 6.0–80. [Internet]. 2018. Available from: https://cran.r-project.org/package=caret

56. Hothorn T, Zeileis A, Cheng E:, Ong S. partykit: A modular toolkit for recursive partytioning in R. J Mach Learn Res. 2015;16:3905–9.

57. Hothorn T, Hornik K, Zeileis A. Unbiased Recursive Partitioning: A Conditional Inference Framework. J Comput Graph Stat [Internet]. Taylor & Francis; 2006 [cited 2018 Nov 24];15:651–74. Available from: http://www.tandfonline.com/doi/abs/10.1198/106186006X133933

58. Fukuyama J, Rumker L, Sankaran K, Jeganathan P, Dethlefsen L, Relman DA, et al. Multidomain analyses of a longitudinal human microbiome intestinal cleanout perturbation experiment. PLoS Comput Biol [Internet]. Public Library of Science; 2017 [cited 2018 Mar 2];13:e1005706. Available from: http://www.ncbi.nlm.nih.gov/pubmed/28821012

59. Njage PMK, Henri C, Leekitcharoenphon P, Mistou M, Hendriksen RS, Hald T. Machine Learning Methods as a Tool for Predicting Risk of Illness Applying Next-Generation Sequencing Data. Risk Anal [Internet]. John Wiley & Sons, Ltd (10.1111); 2018 [cited 2018 Dec 24];risa.13239. Available from: https://onlinelibrary.wiley.com/doi/abs/10.1111/risa.13239

60. Njage PMK, Leekitcharoenphon P, Hald T. Improving hazard characterization in microbial risk assessment using next generation sequencing data and machine learning: Predicting clinical outcomes in shigatoxigenic Escherichia coli. Int J Food Microbiol [Internet]. Elsevier; 2019 [cited 2018 Dec 24];292:72–82. Available from: https://www.sciencedirect.com/science/article/pii/S0168160518308936#f0005

61. Chen J, Zhang L. GMPR: Geometric Mean of Pairwise Ratios. R package version 0.1.3. 2017.

62. Love MI, Huber W, Anders S. Moderated estimation of fold change and dispersion for RNA-seq data with DESeq2. Genome Biol [Internet]. BioMed Central; 2014 [cited 2018 Jan 24];15:550. Available from: http://genomebiology.biomedcentral.com/articles/10.1186/s13059-014-0550-8

63. McMurdie PJ, Holmes S. Waste Not, Want Not: Why Rarefying Microbiome Data Is Inadmissible. McHardy AC, editor. PLoS Comput Biol [Internet]. 2014 [cited 2016 Apr 13];10:e1003531. Available from: http://dx.plos.org/10.1371/journal.pcbi.1003531

64. Kursa MB, Rudnicki WR. Feature Selection with the Boruta Package. J. Stat. Softw. 2010. p. 1–13.

65. Anderson-Bergman C. icenReg : Regression Models for Interval Censored Data in R. J Stat Softw [Internet]. 2017;81. Available from: http://www.jstatsoft.org/v81/i12/

66. Stekhoven DJ, Buhlmann P. MissForest--non-parametric missing value imputation for mixed-type data. Bioinformatics [Internet]. Oxford University Press; 2012 [cited 2018 Sep 18];28:112–8. Available from: https://academic.oup.com/bioinformatics/article-lookup/doi/10.1093/bioinformatics/btr597

67. Lee D, Lee W, Lee Y, Pawitan Y. Sparse partial least-squares regression and its applications to high-throughput data analysis. Chemom Intell Lab Syst [Internet]. Elsevier; 2011 [cited 2018 Jan 24];109:1–8. Available from: https://www.sciencedirect.com/science/article/pii/S016974391100150X

68. Oksanen J, Blanchet FG, Friendly M, Kindt R, Legendre P, McGlinn D, et al. vegan: Community Ecology Package. R package version 2.5-2. [Internet]. 2018. Available from: https://cran.r-project.org/package=vegan

69. Foster ZSL, Sharpton TJ, Grünwald NJ. Metacoder: An R package for visualization and manipulation of community taxonomic diversity data. PLoS Comput Biol. 2017;13:1–15.

70. Cao Y. microbiomeMarker: microbiome biomarker analysis. R package version 0.0.1.9000. https://github.com/yiluheihei/microbiomeMarker. 2021.

