## Additional File 1 for "Microbiota long-term dynamics and prediction of acute graft-versus-host-disease in pediatric allogeneic stem cell transplantation"

### Additional File 1 – Supplementary Tables

**Supplementary Table S1. Patient characteristics.** Abbreviations: HLA, human leukocyte antigen; TBI, total body irradiation; CY, Cyclophosphamide; VP16, Etoposide; BU, Busulfan; MEL, Melphalan; GvHD, graft-versus-host disease.

| **Characteristics** | | **Number of patients** | **Percentage of all patients (%)** |
| --- | --- | --- | --- |
| **Transplant recipients** | | 29 | 100 |
| **Average recipient age in years** | | 9.5 (Range: 2.5-16.4) | NA |
| **Gut microbiota characterized** | **At ≤ 6 timepoints** | 9 | 31 |
|  | **At 7-8 timepoints** | 13 | 44.8 |
|  | **At 9-10 timepoints** | 7 | 24.1 |
| **Oral microbiota chracterized** | **At ≤ 6 timepoints** | 3 | 10.3 |
|  | **At 7-8 timepoints** | 11 | 37.9 |
|  | **At 9-10 timepoints** | 15 | 51.7 |
| **Nasal microbiota chracterized** | **At ≤ 6 timepoints** | 3 | 10.3 |
|  | **At 7-8 timepoints** | 9 | 31 |
|  | **At 9-10 timepoints** | 17 | 58.6 |
| **Patient sex** | **Female** | 13 | 44.8 |
|  | **Male** | 16 | 55.2 |
| **Disease at transplantation** | **Malignant hematologic diseases** | 20 | 69 |
|  | **Severe aplastic anemia** | 1 | 3.4 |
|  | **Other benign disorders including immunodeficiencies** | 8 | 27.6 |
| **Donor type** | **HLA-matched sibling** | 14 | 48.3 |
|  | **HLA-matched unrelated donor (9/10 or 10/10 match)** | 12 | 41.4 |
|  | **Haplo-identical related donor (5/10 match)** | 3 | 10.3 |
| **Stem cell source** | **Bone marrow** | 23 | 79.3 |
|  | **Umbilical cord blood** | 2 | 6.9 |
|  | **Peripheral blood** | 4 | 13.8 |
| **Conditioning regimen** | **TBI + CY / TBI + VP16** | 6 | 20.7 |
|  | **Combinations of BU, CY, VP16 and MEL** | 6 | 20.7 |
|  | **Fludarabine-thiothepa combinations** | 12 | 41.4 |
|  | **Other combinations with fludarabine** | 5 | 17.2 |
| **Anti-thymocyte globulin treatment** | | 16 | 55.2 |
| **Antibiotics pre- and post-HSCT** | | 29 | 100 |
| **Sex mismatch (female donor to male recipient)** | | 7 | 24.1 |
| **Acute GvHD** | **Grade 0-I** | 20 | 69 |
|  | **Grade II-IV** | 9 | 31 |
| **Infections** | **At least one registered bacterial infection** | 25 | 86.2 |
|  | **At least one registered viral infection** | 29 | 100 |
|  | **At least one registered fungal infection** | 14 | 48.3 |
| **Overall Survival*** | **Alive** | 26 | 89.7 |
|  | **Dead** | 3 | 10.3 |
| **Relapse of primary disease*** | | 2 | 6.9 |
| **Non-relapse mortality** | | 2 | 6.9 |
| **Re-transplantation** | | 1 | 3.4 |

*Mean follow-up time after HSCT: 21.4 months (range: 10.1 – 32.7 months)

**Supplementary Table S2. Taxonomy of a subset of LDA clade members and corresponding LDA-coefficients in the gut, oral cavity, and nasal cavity.**

| **ASV number** | **Body site** | **Phylum** | **Family** | **Genus/Species/**  **Description** | **LDA coefficient (beta)** | **Number of patients with presence of this ASV at ≥ 1 time point** |
| --- | --- | --- | --- | --- | --- | --- |
|  | **Gut** |  |  |  |  |  |
| ASV_1 | gut | *Firmicutes* | *Enterococcaceae* | *Enterococcus* sp. | 0.082 | 29 |
| ASV_30 | gut | *Firmicutes* | *Enterococcaceae* | *Enterococcus* sp. | 0.006 | 26 |
| ASV_158 | gut | *Firmicutes* | *Enterococcaceae* | *Enterococcus* sp. | 0.006 | 13 |
| ASV_178 | gut | *Firmicutes* | *Enterococcaceae* | *Enterococcus* sp. | 0.006 | 12 |
| ASV_344 | gut | *Firmicutes* | *Enterococcaceae* | *Enterococcus* sp. | 0.006 | 8 |
| ASV_395 | gut | *Firmicutes* | *Enterococcaceae* | *Enterococcus* sp. | 0.012 | 14 |
| ASV_424 | gut | *Firmicutes* | *Enterococcaceae* | *Enterococcus* sp. | 0.012 | 9 |
| ASV_543 | gut | *Firmicutes* | *Enterococcaceae* | *Enterococcus* sp. | 0.006 | 7 |
| ASV_552 | gut | *Firmicutes* | *Enterococcaceae* | *Enterococcus* *avium* | 0.006 | 7 |
| ASV_720 | gut | *Firmicutes* | *Enterococcaceae* | *Enterococcus* sp. | 0.078 | 15 |
| ASV_730 | gut | *Firmicutes* | *Enterococcaceae* | *Enterococcus* sp. | 0.006 | 12 |
| ASV_784 | gut | *Firmicutes* | *Enterococcaceae* | *Enterococcus* sp. | 0.006 | 8 |
| ASV_951 | gut | *Firmicutes* | *Enterococcaceae* | *Enterococcus* sp. | 0.006 | 12 |
| ASV_1186 | gut | *Firmicutes* | *Enterococcaceae* | *Melissococcus* sp. | 0.006 | 9 |
| ASV_3 | gut | *Firmicutes* | *Lactobacillaceae* | *Lactobacillus* sp. | 0.011 | 26 |
| ASV_31 | gut | *Firmicutes* | *Lactobacillaceae* | *Lactobacillus* sp. | 0.006 | 11 |
| ASV_67 | gut | *Firmicutes* | *Lactobacillaceae* | *Lactobacillus sakei* | 0.006 | 15 |
| ASV_74 | gut | *Firmicutes* | *Lactobacillaceae* | *Lactobacillus* sp. | 0.006 | 16 |
| ASV_144 | gut | *Firmicutes* | *Lactobacillaceae* | *Pediococcus* sp. | 0.006 | 10 |
| ASV_151 | gut | *Firmicutes* | *Lactobacillaceae* | *Pediococcus pentosaceus* | 0.006 | 12 |
| ASV_189 | gut | *Firmicutes* | *Lactobacillaceae* | *Lactobacillus* sp. | 0.006 | 10 |
| ASV_222 | gut | *Firmicutes* | *Lactobacillaceae* | *Lactobacillus rhamnosus* | 0.006 | 8 |
| ASV_586 | gut | *Firmicutes* | *Lactobacillaceae* | *Lactobacillus* sp. | 0.006 | 9 |
| ASV_78 | gut | *Firmicutes* | *Lachnospiraceae* | *Blautia* sp. | -0.039 | 26 |
| ASV_237 | gut | *Firmicutes* | *Lachnospiraceae* | *Blautia faecis* | -0.026 | 23 |
| ASV_265 | gut | *Firmicutes* | *Lachnospiraceae* | *Blautia* sp. | -0.026 | 24 |
| ASV_271 | gut | *Firmicutes* | *Lachnospiraceae* | *Blautia obeum* | -0.026 | 12 |
| ASV_375 | gut | *Firmicutes* | *Lachnospiraceae* | *Blautia* sp. | -0.026 | 15 |
| ASV_554 | gut | *Firmicutes* | *Lachnospiraceae* | *Blautia* sp. | -0.026 | 16 |
| ASV_1291 | gut | *Firmicutes* | *Lachnospiraceae* | *Blautia* sp. | -0.026 | 9 |
| ASV_132 | gut | *Firmicutes* | *Ruminococcaceae* | *Ruminococcus bromii* | -0.007 | 16 |
| ASV_192 | gut | *Firmicutes* | *Ruminococcaceae* | *Ruminococcus bromii* | -0.007 | 24 |
| ASV_206 | gut | *Firmicutes* | *Ruminococcaceae* | DTU089 | -0.024 | 23 |
| ASV_306 | gut | *Firmicutes* | *Ruminococcaceae* | *Ruminococcus* sp. | -0.007 | 13 |
| ASV_348 | gut | *Firmicutes* | *Ruminococcaceae* | *Ruminococcus* sp. | -0.007 | 11 |
| ASV_556 | gut | *Firmicutes* | *Ruminococcaceae* | *Caproiciproducens* sp. | -0.024 | 9 |
| ASV_566 | gut | *Firmicutes* | *Ruminococcaceae* | *Ruminoclostridium sp.* | -0.024 | 13 |
| ASV_583 | gut | *Firmicutes* | *Ruminococcaceae* | *Ruminoclostridium sp.* | -0.024 | 23 |
| ASV_682 | gut | *Firmicutes* | *Ruminococcaceae* | *Caproiciproducens* sp. | -0.024 | 9 |
| ASV_750 | gut | *Firmicutes* | *Ruminococcaceae* | *Caproiciproducens* sp. | -0.024 | 13 |
| ASV_792 | gut | *Firmicutes* | *Ruminococcaceae* | *Ruminoclostridium sp.* | -0.024 | 15 |
|  | **Oral** |  |  |  |  |  |
| ASV_18 | oral | *Actinobacteria* | *Actinomycetaceae* | *Actinomyces* sp. | 0.021 | 27 |
| ASV_66 | oral | *Actinobacteria* | *Actinomycetaceae* | *Actinomyces* sp. | 0.003 | 26 |
| ASV_117 | oral | *Actinobacteria* | *Actinomycetaceae* | *Actinomyces naeslundii* | 0.016 | 27 |
| ASV_126 | oral | *Actinobacteria* | *Actinomycetaceae* | *Actinomyces naeslundii* | 0.016 | 19 |
| ASV_138 | oral | *Actinobacteria* | *Actinomycetaceae* | *Actinomyces* sp. | 0.003 | 22 |
| ASV_155 | oral | *Actinobacteria* | *Actinomycetaceae* | *Actinomyces odontolyticus* | 0.003 | 20 |
| ASV_227 | oral | *Actinobacteria* | *Actinomycetaceae* | *Actinomyces* sp. | 0.016 | 7 |
| ASV_235 | oral | *Actinobacteria* | *Actinomycetaceae* | F0332 | 0.003 | 18 |
| ASV_262 | oral | *Actinobacteria* | *Actinomycetaceae* | *Actinomyces* sp. | 0.003 | 18 |
| ASV_345 | oral | *Actinobacteria* | *Actinomycetaceae* | *Actinomyces* sp. | 0.016 | 9 |
| ASV_389 | oral | *Actinobacteria* | *Actinomycetaceae* | *Actinomyces odontolyticus* | 0.003 | 7 |
| ASV_403 | oral | *Actinobacteria* | *Actinomycetaceae* | *Actinomyces* sp. | 0.003 | 15 |
| ASV_407 | oral | *Actinobacteria* | *Actinomycetaceae* | *Actinomyces* sp. | 0.016 | 7 |
| ASV_2693 | oral | *Actinobacteria* | *Actinomycetaceae* | *Actinomyces* sp. | 0.021 | 5 |
| ASV_422 | oral | *Actinobacteria* | *Actinomycetaceae* | *Actinomyces odontolyticus* | 0.003 | 15 |
| ASV_431 | oral | *Actinobacteria* | *Actinomycetaceae* | *Actinomyces* sp. | 0.016 | 12 |
| ASV_2697 | oral | *Actinobacteria* | *Actinomycetaceae* | *Actinomyces naeslundii* | 0.016 | 9 |
| ASV_461 | oral | *Actinobacteria* | *Actinomycetaceae* | *Actinomyces* sp. | 0.003 | 11 |
| ASV_475 | oral | *Actinobacteria* | *Actinomycetaceae* | *Actinomyces gerencseriae* | 0.003 | 6 |
| ASV_484 | oral | *Actinobacteria* | *Actinomycetaceae* | *Actinomyces* sp. | 0.003 | 4 |
| ASV_501 | oral | *Actinobacteria* | *Actinomycetaceae* | *Actinomyces odontolyticus* | 0.003 | 13 |
| ASV_516 | oral | *Actinobacteria* | *Actinomycetaceae* | *Actinomyces odontolyticus* | 0.003 | 12 |
| ASV_2700 | oral | *Actinobacteria* | *Actinomycetaceae* | *Actinomyces gerencseriae* | 0.003 | 14 |
| ASV_568 | oral | *Actinobacteria* | *Actinomycetaceae* | *Actinomyces* sp. | 0.003 | 12 |
| ASV_600 | oral | *Actinobacteria* | *Actinomycetaceae* | *Actinomyces* sp. | 0.003 | 7 |
| ASV_642 | oral | *Actinobacteria* | *Actinomycetaceae* | *Actinomyces* sp. | 0.003 | 7 |
| ASV_798 | oral | *Actinobacteria* | *Actinomycetaceae* | F0332 | 0.003 | 10 |
| ASV_871 | oral | *Actinobacteria* | *Actinomycetaceae* | *Actinomyces massiliensis* | 0.003 | 13 |
| ASV_2729 | oral | *Actinobacteria* | *Actinomycetaceae* | *Actinomyces graevenitzii* | 0.003 | 9 |
| ASV_1055 | oral | *Actinobacteria* | *Actinomycetaceae* | *Actinomyces* sp. | 0.003 | 6 |
| ASV_1172 | oral | *Actinobacteria* | *Actinomycetaceae* | *Actinomyces* sp. | 0.003 | 7 |
| ASV_2751 | oral | *Actinobacteria* | *Actinomycetaceae* | *Actinomyces* sp. | 0.016 | 8 |
| ASV_2664 | oral | *Firmicutes* | *Streptococcaceae* | *Streptococcus* sp. | 0.010 | 29 |
| ASV_10 | oral | *Firmicutes* | *Streptococcaceae* | *Streptococcus* sp. | 0.010 | 29 |
| ASV_16 | oral | *Firmicutes* | *Streptococcaceae* | *Streptococcus* sp. | 0.010 | 29 |
| ASV_27 | oral | *Firmicutes* | *Streptococcaceae* | *Streptococcus* sp. | 0.010 | 29 |
| ASV_28 | oral | *Firmicutes* | *Streptococcaceae* | *Streptococcus* sp. | 0.040 | 28 |
| ASV_37 | oral | *Firmicutes* | *Streptococcaceae* | *Streptococcus* sp. | 0.010 | 19 |
| ASV_48 | oral | *Firmicutes* | *Streptococcaceae* | *Streptococcus* sp. | 0.010 | 27 |
| ASV_173 | oral | *Firmicutes* | *Streptococcaceae* | *Streptococcus salivarius* | 0.010 | 15 |
| ASV_183 | oral | *Firmicutes* | *Streptococcaceae* | *Streptococcus* sp. | 0.010 | 19 |
| ASV_188 | oral | *Firmicutes* | *Streptococcaceae* | *Streptococcus* sp. | 0.010 | 14 |
| ASV_2674 | oral | *Firmicutes* | *Streptococcaceae* | *Streptococcus* sp. | 0.010 | 14 |
| ASV_230 | oral | *Firmicutes* | *Streptococcaceae* | *Streptococcus* sp. | 0.010 | 8 |
| ASV_269 | oral | *Firmicutes* | *Streptococcaceae* | *Streptococcus cristatus* | 0.010 | 29 |
| ASV_282 | oral | *Firmicutes* | *Streptococcaceae* | *Streptococcus parasanguinis* | 0.010 | 13 |
| ASV_2683 | oral | *Firmicutes* | *Streptococcaceae* | *Streptococcus mitis* | 0.010 | 11 |
| ASV_480 | oral | *Firmicutes* | *Streptococcaceae* | *Streptococcus* sp. | 0.010 | 18 |
| ASV_481 | oral | *Firmicutes* | *Streptococcaceae* | *Streptococcus peroris* | 0.010 | 12 |
| ASV_802 | oral | *Firmicutes* | *Streptococcaceae* | *Streptococcus* sp. | 0.010 | 24 |
| ASV_1531 | oral | *Firmicutes* | *Streptococcaceae* | *Streptococcus* sp. | 0.010 | 11 |
| ASV_1599 | oral | *Firmicutes* | *Streptococcaceae* | *Streptococcus* sp. | 0.010 | 13 |
| ASV_42 | oral | *Bacteroidetes* | *Prevotellaceae* | *Prevotella melaninogenica* | 0.028 | 27 |
| ASV_226 | oral | *Bacteroidetes* | *Prevotellaceae* | *Prevotella melaninogenica* | 0.028 | 15 |
| ASV_800 | oral | *Bacteroidetes* | *Prevotellaceae* | *Prevotella* sp. | 0.028 | 6 |
| ASV_2665 | oral | *Firmicutes* | Family XI | *Gemella* sp. | 0.009 | 29 |
| ASV_208 | oral | *Firmicutes* | Family XI | *Gemella sanguinis* | 0.009 | 28 |
| ASV_2701 | oral | *Firmicutes* | Family XI | *Gemella* sp. | 0.009 | 5 |
|  | **Nasal** |  |  |  |  |  |
| ASV_14 | nasal | *Actinobacteria* | *Corynebacteriaceae* | *Corynebacterium* sp. | 0.117 | 24 |
| ASV_354 | nasal | *Actinobacteria* | *Corynebacteriaceae* | *Corynebacterium durum* | 0.054 | 15 |
| ASV_360 | nasal | *Actinobacteria* | *Corynebacteriaceae* | *Corynebacterium* sp. | 0.054 | 17 |
| ASV_2704 | nasal | *Actinobacteria* | *Corynebacteriaceae* | *Corynebacterium* sp. | 0.022 | 12 |
| ASV_2707 | nasal | *Actinobacteria* | *Corynebacteriaceae* | *Corynebacterium durum* | 0.054 | 14 |
| ASV_1206 | nasal | *Actinobacteria* | *Corynebacteriaceae* | *Lawsonella* sp. | 0.007 | 16 |

**Supplementary Table S3. Taxonomy of aGvHD predictors within the fecal, oral, and nasal microbiota.** ASVs that were significantly predicting aGvHD severity according to the conditional inference tree regression model are highlighted in bold. Of the 50 most important gut ASVs identified by the svmLinear model, 17 were confirmed by Boruta feature selection and are listed here. In the oral and nasal cavities, 26 and 12 ASVs were confirmed by Boruta selection, respectively. Listed in bold are those ASVs with a significant predictive effect on aGvHD severity, tested in a regression framework with CTREE (see Methods).

| **ASV number** | **Phylum** | **Family** | **Genus/Species/Description** | **Body Site** | **Importance in svmLinear model** | **Mean Importance in Boruta** |
| --- | --- | --- | --- | --- | --- | --- |
|  | **Gut** |  |  |  |  |  |
| **ASV_3** | ***Firmicutes*** | ***Lactobacillaceae*** | ***Lactobacillus sp.*** | **gut** | **0.74** | **8.85** |
| ASV_7 | *Bacteroidetes* | *Tannerellaceae* | *Parabacteroides merdae* | gut | 0.63 | 4.62 |
| ASV_8 | *Proteobacteria* | *Enterobacteriaceae* | *Escherichia/Shigella* sp. | gut | 0.70 | 4.16 |
| ASV_12 | *Firmicutes* | *Ruminococcaceae* | UBA1819 sp. | gut | 0.68 | 5.52 |
| ASV_35 | *Bacteroidetes* | *Bacteroidaceae* | *Bacteroides uniformis* | gut | 0.68 | 4.66 |
| ASV_50 | *Firmicutes* | *Ruminococcaceae* | *Subdoligranulum* sp. | gut | 0.60 | 3.78 |
| **ASV_128** | ***Bacteroidetes*** | ***Tannerellaceae*** | ***Parabacteroides distasonis*** | **gut** | **0.68** | **7.42** |
| ASV_131 | *Bacteroidetes* | *Tannerellaceae* | *Parabacteroides distasonis* | gut | 0.66 | 6.49 |
| ASV_166 | *Fusobacteria* | *Fusobacteriaceae* | *Fusobacterium* sp*.* | gut | 0.60 | 4.08 |
| **ASV_268** | ***Firmicutes*** | ***Lachnospiraceae*** | ***Lachnospiraceae_NK4A136_group* sp.** | **gut** | **0.61** | **5.02** |
| ASV_361 | *Bacteroidetes* | *Tannerellaceae* | *Parabacteroides* sp. | gut | 0.61 | 4.40 |
| ASV_477 | *Firmicutes* | *Lachnospiraceae* | *Eisenbergiella tayi* | gut | 0.62 | 3.64 |
| ASV_507 | *Firmicutes* | *Lachnospiraceae* | *Lachnospira pectinoschiza* | gut | 0.60 | 3.91 |
| ASV_563 | *Firmicutes* | *Erysipelotrichaceae* | *Coprobacillus cateniformis* | gut | 0.63 | 5.38 |
| ASV_567 | *Firmicutes* | *Ruminococcaceae* | *Flavonifractor* sp. | gut | 0.60 | 2.81 |
| ASV_585 | *Bacteroidetes* | *Tannerellaceae* | *Parabacteroides* sp. | gut | 0.62 | 4.53 |
| ASV_687 | *Firmicutes* | *Lachnospiraceae* | NA | gut | 0.59 | 3.83 |
|  | **Oral** |  |  |  |  |  |
| ASV_15 | *Firmicutes* | *Streptococcaceae* | *Lactococcus* sp*.* | oral | 0.61 | 7.31 |
| ASV_34 | *Proteobacteria* | *Neisseriaceae* | *Neisseria* sp. | oral | 0.62 | 5.08 |
| ASV_66 | *Actinobacteria* | *Actinomycetaceae* | *Actinomyces* sp. | oral | 0.65 | 4.84 |
| ASV_72 | *Actinobacteria* | *Micrococcaceae* | *Rothia* sp. | oral | 0.63 | 4.49 |
| ASV_92 | *Bacteroidetes* | *Bacteroidaceae* | *Bacteroides* sp. | oral | 0.65 | 5.36 |
| ASV_98 | *Actinobacteria* | *Micrococcaceae* | *Rothia mucilaginosa* | oral | 0.58 | 5.87 |
| ASV_135 | *Firmicutes* | *Streptococcaceae* | *Streptococcus mutans* | oral | 0.60 | 4.24 |
| ASV_205 | *Firmicutes* | *Ruminococcaceae* | *Ruminococcaceae_UCG-002* sp. | oral | 0.63 | 4.38 |
| **ASV_226** | ***Bacteroidetes*** | ***Prevotellaceae*** | ***Prevotella_7 melaninogenica*** | **oral** | **0.63** | **8.19** |
| ASV_247 | *Firmicutes* | *Ruminococcaceae* | *Ruminococcaceae_UCG-013* sp. | oral | 0.66 | 4.80 |
| ASV_270 | *Firmicutes* | *Veillonellaceae* | *Veillonella* sp. | oral | 0.59 | 3.47 |
| ASV_338 | *Bacteroidetes* | *Prevotellaceae* | *Alloprevotella* sp. | oral | 0.61 | 4.26 |
| ASV_360 | *Actinobacteria* | *Corynebacteriaceae* | *Corynebacterium* sp. | oral | 0.64 | 6.50 |
| ASV_374 | *Fusobacteria* | *Leptotrichiaceae* | *Leptotrichia wadei* | oral | 0.60 | 4.39 |
| ASV_378 | *Firmicutes* | *Lachnospiraceae* | *Oribacterium sinus* | oral | 0.65 | 7.17 |
| **ASV_500** | ***Actinobacteria*** | ***Propionibacteriaceae*** | ***Pseudopropionibacterium propionicum*** | **oral** | **0.67** | **8.51** |
| ASV_518 | *Bacteroidetes* | *Bacteroidaceae* | *Bacteroides* sp. | oral | 0.64 | 4.67 |
| **ASV_568** | ***Actinobacteria*** | ***Actinomycetaceae*** | ***Actinomyces* sp.** | **oral** | **0.71** | **8.79** |
| ASV_640 | *Bacteroidetes* | *Tannerellaceae* | *Parabacteroides* sp. | oral | 0.62 | 2.81 |
| ASV_871 | *Actinobacteria* | *Actinomycetaceae* | *Actinomyces massiliensis* | oral | 0.61 | 4.65 |
| ASV_894 | *Firmicutes* | *Ruminococcaceae* | *Ruminococcaceae_UCG-014* sp. | oral | 0.62 | 2.74 |
| ASV_1403 | *Firmicutes* | *Veillonellaceae* | *Selenomonas_3* sp. | oral | 0.59 | 2.82 |
| ASV_2664 | *Firmicutes* | *Streptococcaceae* | *Streptococcus* sp. | oral | 0.68 | 6.11 |
| ASV_2666 | *Actinobacteria* | *Micrococcaceae* | *Rothia dentocariosa* | oral | 0.61 | 4.47 |
| ASV_2676 | *Firmicutes* | *Aerococcaceae* | *Abiotrophia defectiva* | oral | 0.61 | 5.74 |
| ASV_2692 | *Actinobacteria* | *Micrococcaceae* | *Rothia* sp. | oral | 0.62 | 6.30 |
|  | **Nasal** |  |  |  |  |  |
| ASV_2664 | *Firmicutes* | *Streptococcaceae* | *Streptococcus* sp. | nasal | 0.61 | 11.49 |
| ASV_2666 | *Actinobacteria* | *Micrococcaceae* | *Rothia dentocariosa* | nasal | 0.56 | 5.80 |
| **ASV_47** | ***Actinobacteria*** | ***Micrococcaceae*** | ***Rothia* sp.** | **nasal** | **0.59** | **6.41** |
| ASV_51 | *Proteobacteria* | *Pasteurellaceae* | *Haemophilus* sp. | nasal | 0.64 | 5.67 |
| ASV_52 | *Actinobacteria* | *Micrococcaceae* | *Rothia mucilaginosa* | nasal | 0.61 | 8.58 |
| **ASV_66** | ***Actinobacteria*** | ***Actinomycetaceae*** | ***Actinomyces* sp.** | **nasal** | **0.67** | **9.05** |
| ASV_2670 | *Proteobacteria* | *Neisseriaceae* | NA | nasal | 0.61 | 6.69 |
| ASV_117 | *Actinobacteria* | *Actinomycetaceae* | *Actinomyces naeslundii* | nasal | 0.58 | 4.73 |
| ASV_125 | *Actinobacteria* | *Corynebacteriaceae* | *Corynebacterium_1 accolens* | nasal | 0.57 | 5.43 |
| ASV_252 | *Firmicutes* | *Veillonellaceae* | *Veillonella rogosae* | nasal | 0.58 | 5.54 |
| ASV_270 | *Firmicutes* | *Veillonellaceae* | *Veillonella* sp. | nasal | 0.57 | 5.41 |
| ASV_2694 | *Actinobacteria* | *Corynebacteriaceae* | *Lawsonella* sp. | nasal | 0.65 | 8.42 |
