## Additional File 2 for "Microbiota long-term dynamics and prediction of acute graft-versus-host-disease in pediatric allogeneic stem cell transplantation"

### Additional File 2 – Supplementary Figures

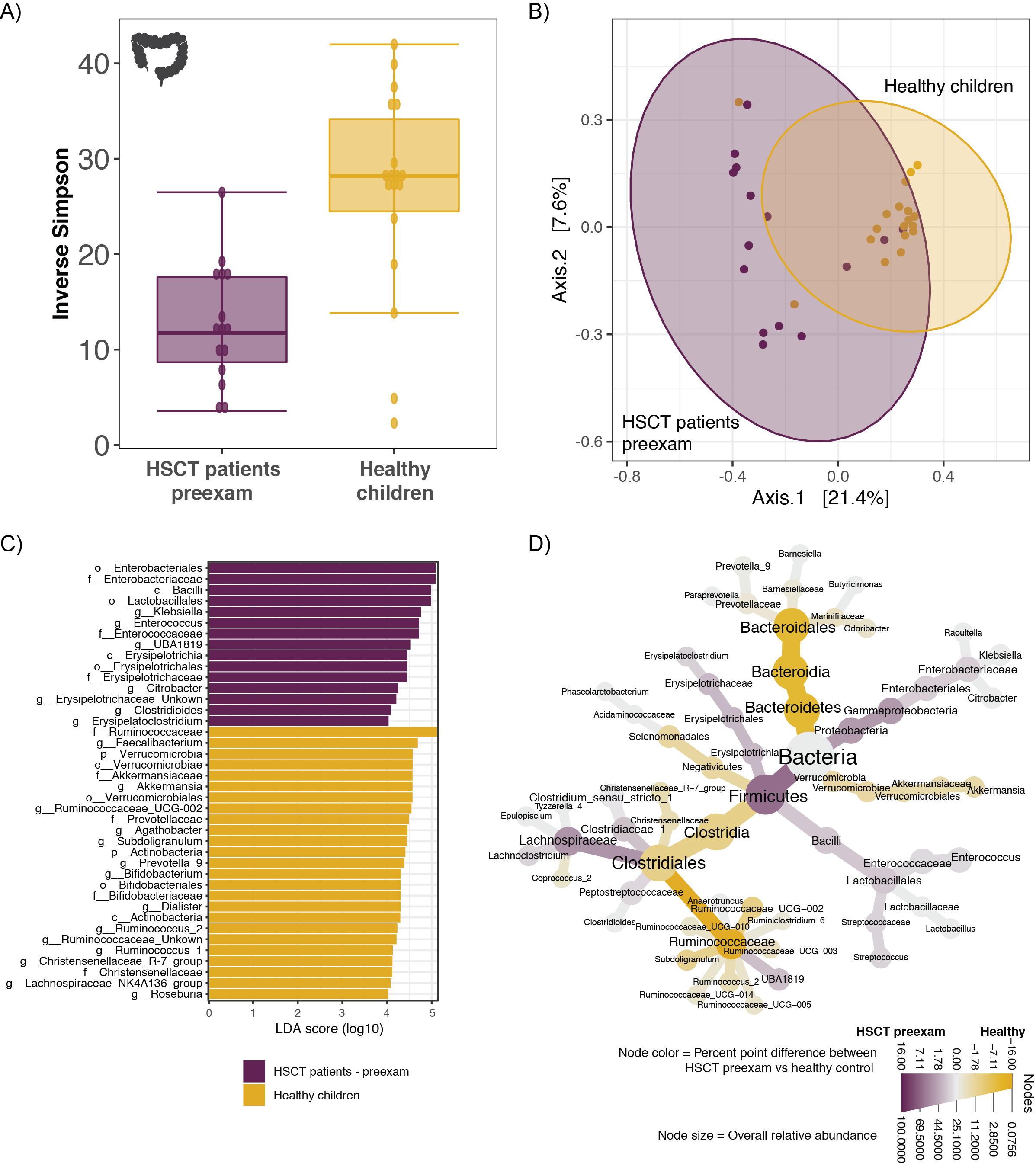

**Figure S1. The gut microbiota in the HSCT patients at pre-exam differs from the gut microbiota of age-matched healthy children.** A) Fecal bacterial alpha diversity (inverse Simpson index) was 2.4-fold higher in healthy children (n=18) compared to children at pre-examination before HSCT (n=15). B) Fecal bacterial composition was significantly different between the two groups (anosim, p=0.001, R=0.44), and within-group variance was significantly greater in the HSCT group (betadisper, p<0.001). C) The taxa which best explain differences in community structure between HSCT patients at preexamination and healthy children were identified by analysis of LEfSe (Linear discriminant analysis Effect Size). LefSe accounts for the hierarchical structure of bacterial phylogeny, thereby allowing identification of differentially abundant taxa on several taxonomic levels (here: kingdom to genus). Count data was centered-log ratio (CLR) transformed within the LEfSe analysis. The higher the LDA score (log10), the higher the effect size of the respective taxon in explaining group difference. Here, we show taxa with an LDA score >4. D) Differentially abundant genera between the two groups were additionally identified by DESeq2. Of the top 100 most abundant genera (of the whole gut microbiota data set), eighteen genera were significantly more abundant in healthy children (yellow), and 15 genera were significantly more abundant in the patients at preexam (purple). Differences in median proportions of these genera (and their supertaxa) are displayed in a heat tree. See also additional information at <https://doi.org/10.6084/m9.figshare.13614230>.

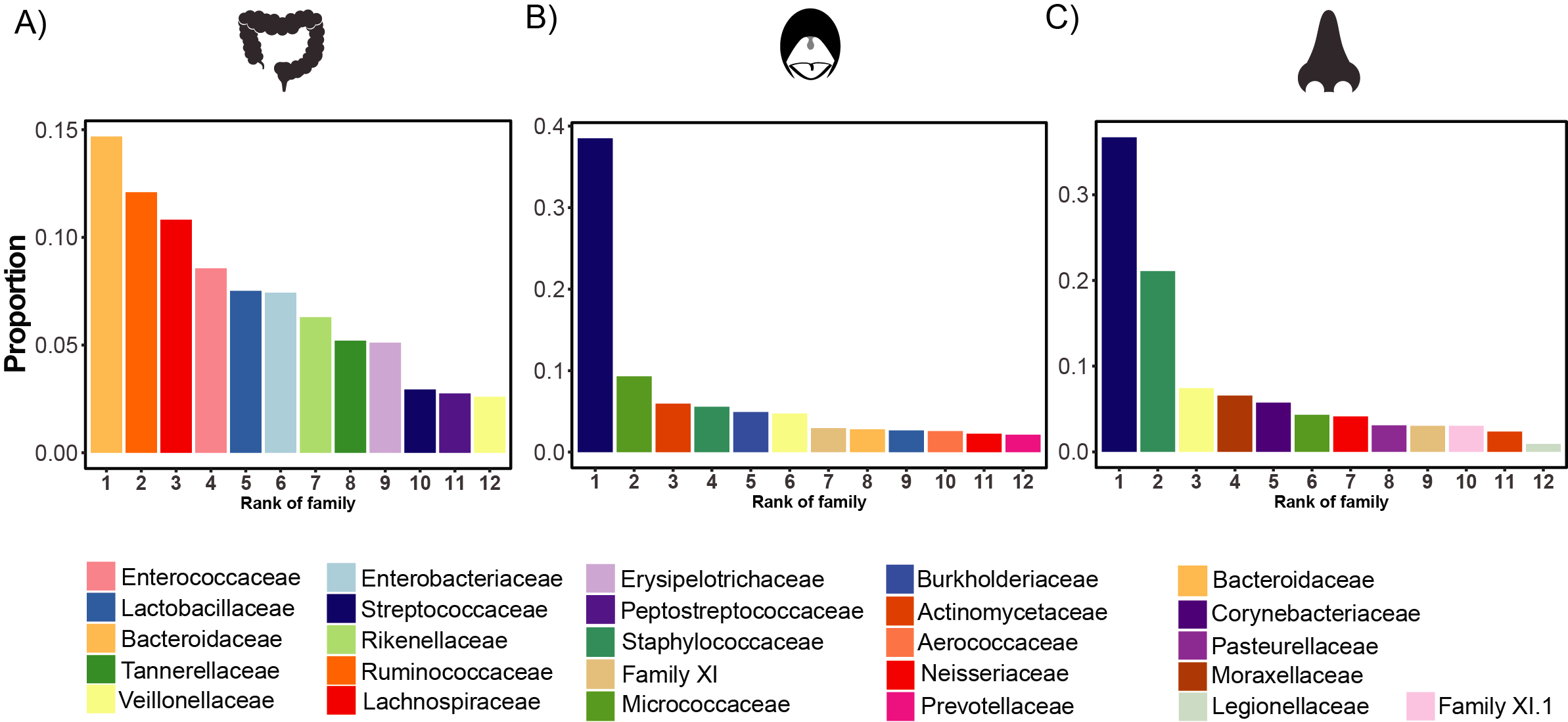

**Figure S2. Most abundant taxonomic families in the gut, oral cavity, and nasal cavity in allo-HSCT patients.** Rank abundance curves displaying the proportions of the 12 most abundant taxonomic families at each body site (gut, oral cavity, and nasal cavity).

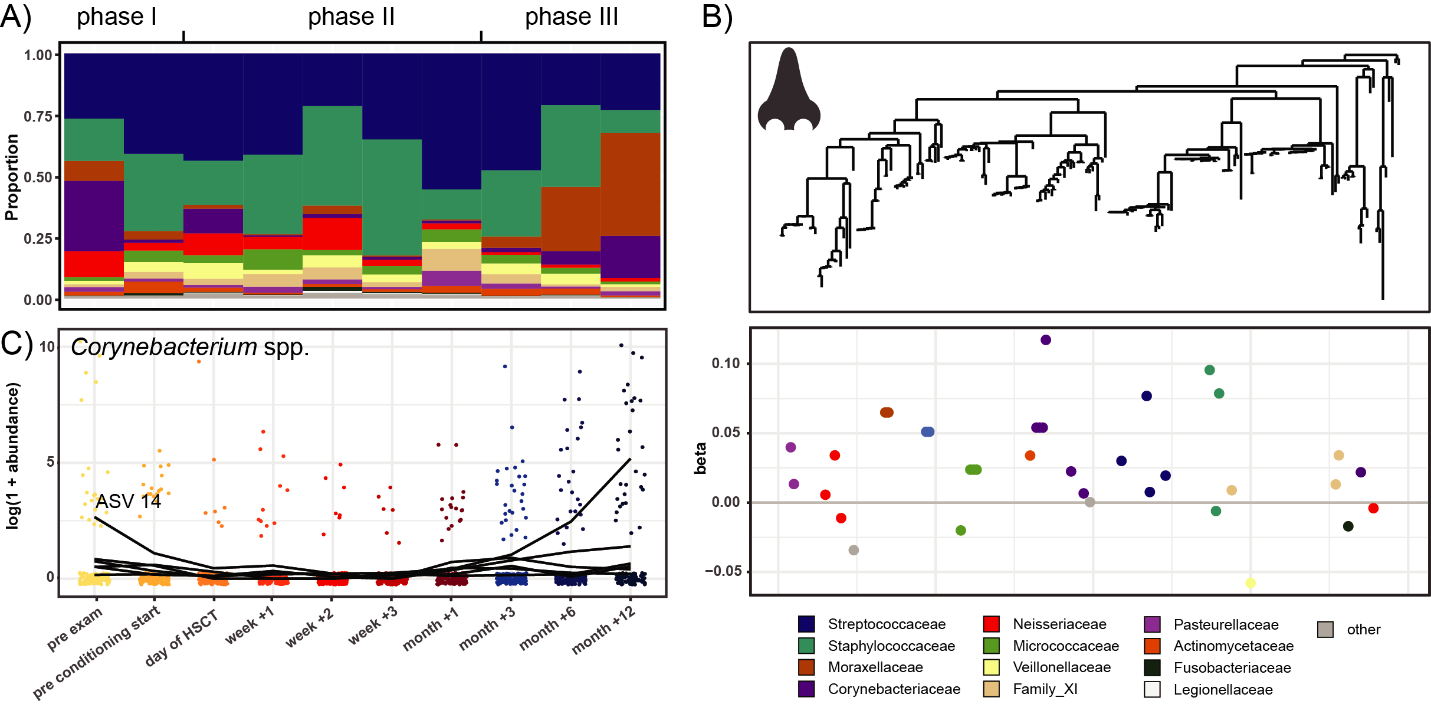

**Figure S3.** **Tree-based sparse linear discriminant analysis revealing nasal ASVs that distinguish time points from each other in relation to HSCT.** A) Relative abundances over time of the 12 most abundant families in the nasal cavity. B) Coefficients of discriminating clades of ASVs on the first LDA axis, colored by taxonomic family, and plotted along the phylogenetic tree. C) Trajectories of ASVs in one discriminating group, affiliated with the family *Corynebacteriaceae,* with decreasing abundances after HSCT and recovery at late follow-up time points. The most abundant discriminating ASV is indicated Detailed taxonomic information and LDA-coefficients of the displayed ASVs are listed in Table S2.

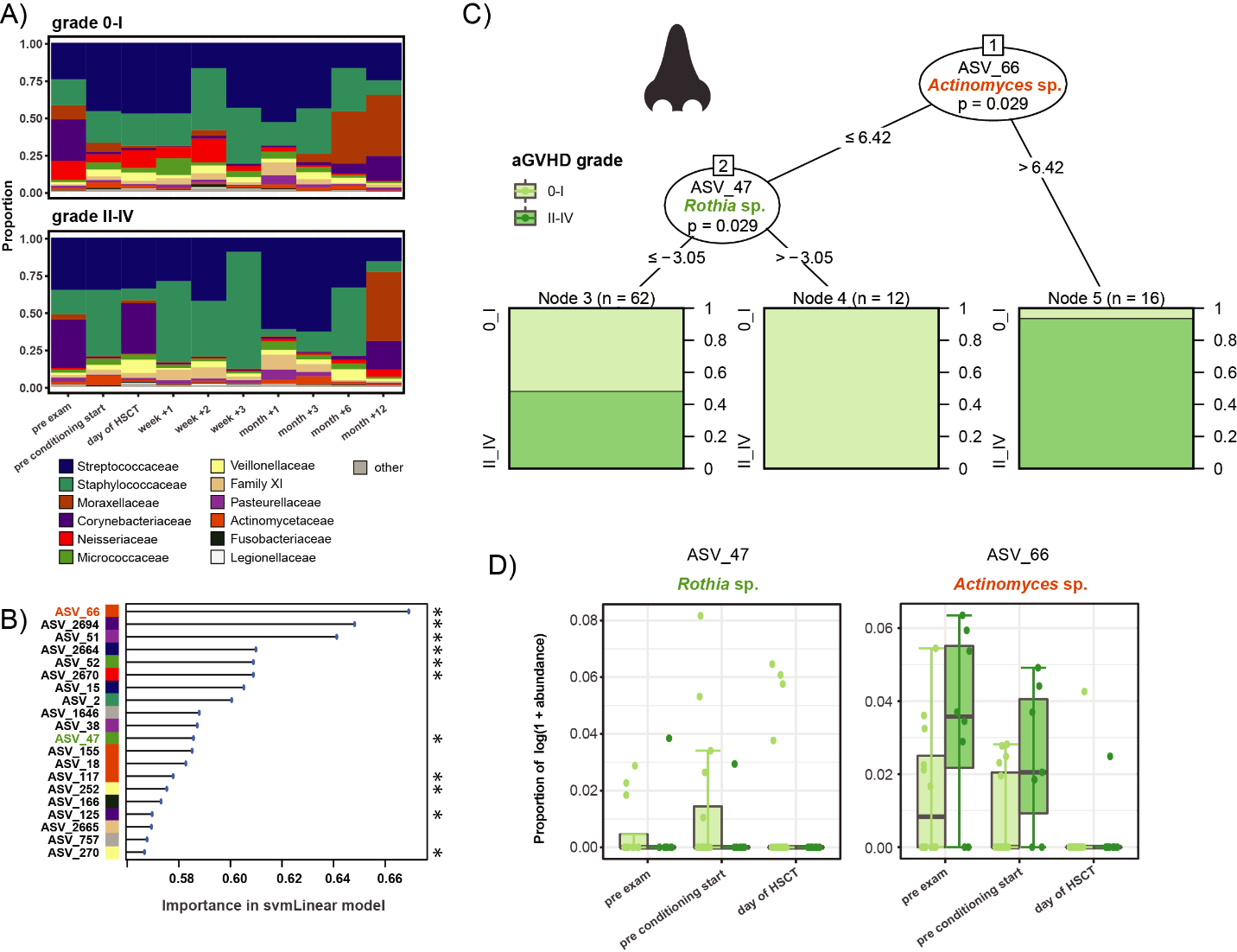

**Figure S4.** **Machine learning-based prediction of aGvHD severity from nasal microbial abundances pre-HSCT.** A) Relative abundances of the 12 most abundant families over time in the nasal cavity in patients with aGvHD grade 0-I versus II-IV. B) Importance plot of top 20 predictive nasal ASVs identified by the svmLinear model with importance scores indicating the mean decrease in prediction accuracy in case the respective ASV would be excluded from the model. The final cross-validated svmLinear model predicted aGvHD (0-I versus II-IV) from the abundances of nasal ASVs pre-HSCT with 76% accuracy (95% CI: 56% to 90%). The ASVs that were also confirmed by Boruta feature selection are indicated with asterisk. C) Conditional inference tree (CTREE) displaying ASVs identified as significant split nodes by nonparametric regression for prediction of aGvHD. Numbers along the branches indicate split values of variance stabilized bacterial abundances. The terminal nodes show the proportion of samples originating from patients with aGvHD grade 0-I vs II-IV (n = number of samples). D) Boxplots depict the log transformed relative abundances of the predictive ASVs at time points up to the transplantation in aGvHD grade 0-I compared with grade II-IV patients.

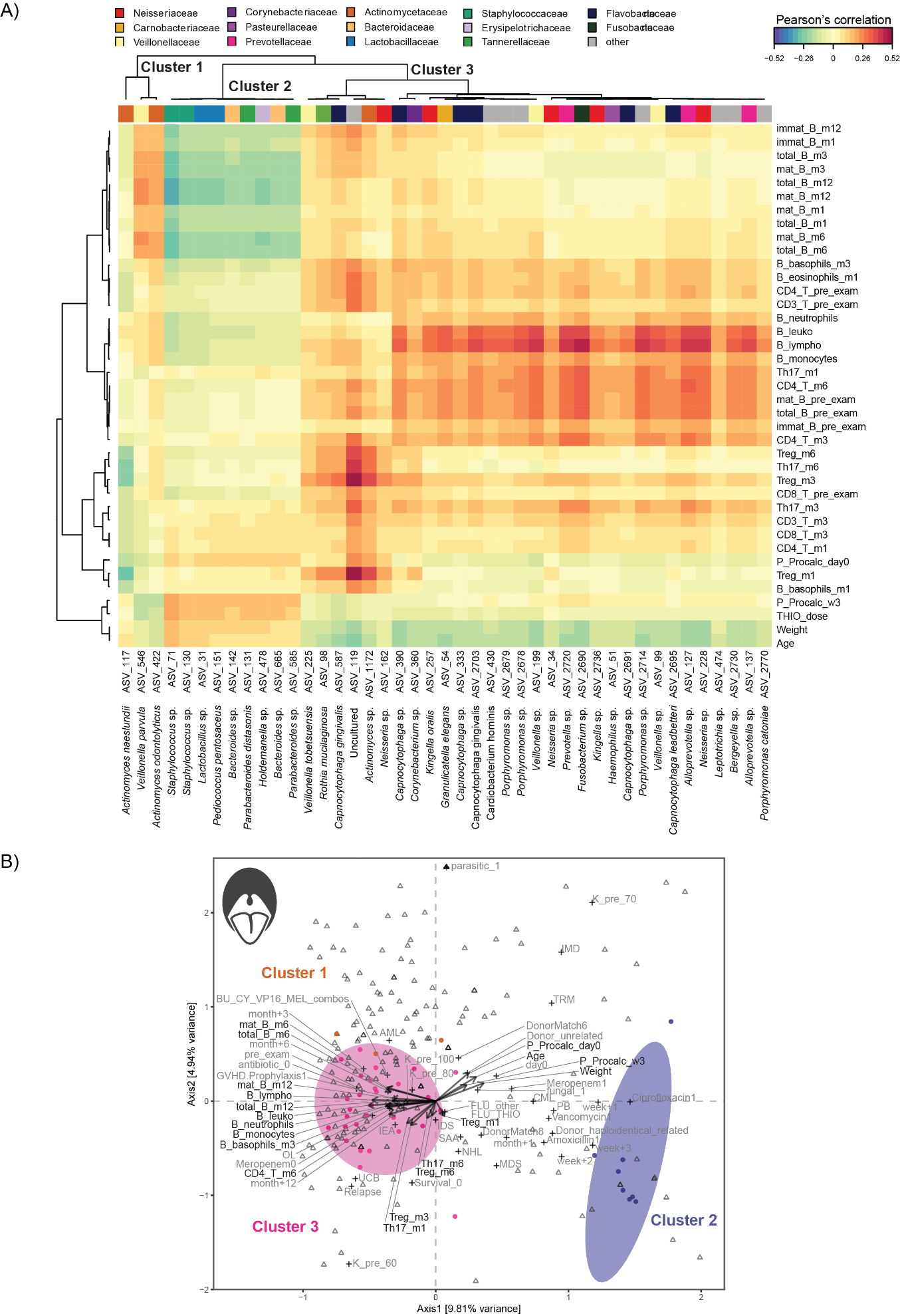

**Figure S5. Multivariate associations of the oral microbiota with immune and clinical parameters in HSCT.** A) Clustered image map (CIM) based on sparse partial least squares (sPLS) regression analysis dimensions 1, 2, and 3, displaying pairwise correlations >0.2/<-0.2 between oral ASVs (bottom), and continuous immune and clinical parameters (right). Red indicated positive correlation, and blue indicates negative correlation, respectively. Based on the sPLS regression model, hierarchical clustering (clustering method: complete linkage, distance method: Pearson’s correlation) was performed resulting in the three depicted clusters. B) Canonical correspondence analysis (CCpnA) relating oral microbial abundances (circles) to continuous (arrows) and categorical (+) immune and clinical parameters. ASVs and variables with at least one correlation >0.2/<-0.2 in the sPLS analysis were included in the CCpnA. The triplot shows variables and ASVs with a score >0.3/<-0.3 on at least one the first three CCpnA axes, displayed on axis 1 versus 2 with samples depicted as triangles. The colored ellipses (depicted with 80% confidence interval) correspond to the clusters of ASVs identified by the sPLS-based hierarchical clustering. For visualization purposes, a focused section of the CCpnA triplot is shown. Abbreviations are described in Figure 6. Additional abbreviations: fungal, fungal infection; haploident, haploidentical donor; hemo, hemoglobin; leuko, leukocytes; lympho, lymphocytes; w1, week+1; w2, week+2; w3, week+3.

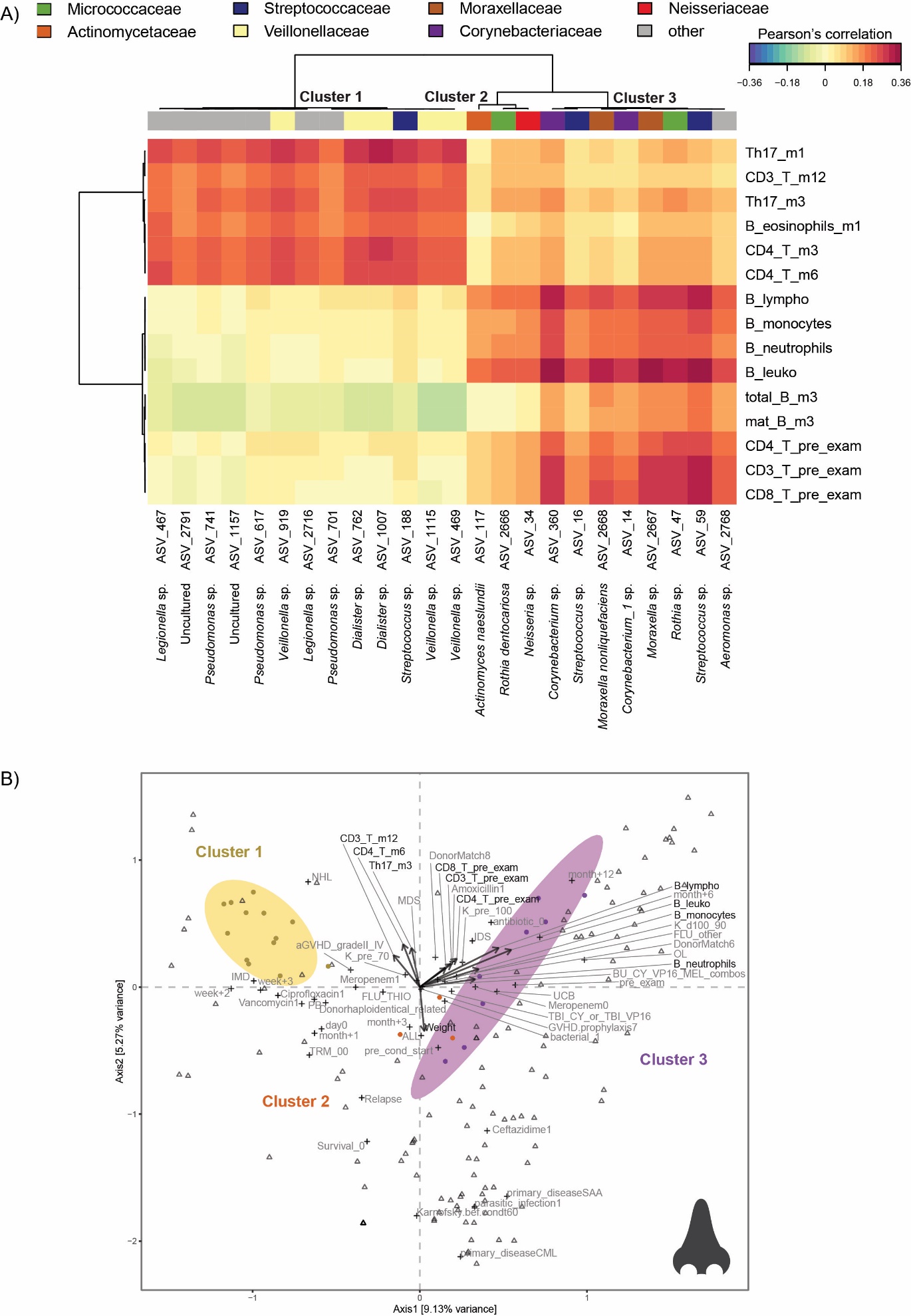

**Figure S6. Multivariate associations of the nasal microbiota with immune and clinical parameters in HSCT.** A) Clustered image map (CIM) based on sparse partial least squares (sPLS) regression analysis dimensions 1, 2, and 3, displaying pairwise correlations >0.2/<-0.2 between nasal ASVs (bottom), and continuous immune and clinical parameters (right). Red indicated positive correlation, and blue indicates negative correlation, respectively. Based on the sPLS regression model, hierarchical clustering (clustering method: complete linkage, distance method: Pearson’s correlation) was performed resulting in the three depicted clusters. B) Canonical correspondence analysis (CCpnA) relating nasal microbial abundances (circles) to continuous (arrows) and categorical (+) immune and clinical parameters. ASVs and variables with at least one correlation >0.2/<-0.2 in the sPLS analysis were included in the CCpnA. The triplot shows variables and ASVs with a score >0.3/<-0.3 on at least one the first three CCpnA axes, displayed on axis 1 versus 2 with samples depicted as triangles. The colored ellipses (depicted with 80% confidence interval) correspond to the clusters of ASVs identified by the sPLS-based hierarchical clustering. For visualization purposes, a focused section of the CCpnA triplot is shown. Abbreviations are described in Figures 6 and S5. Additional abbreviations: DonorMatch8, unrelated donor with 1 HLA mismatch; PB, peripheral blood.
