## Additional File 3 for "Microbiota long-term dynamics and prediction of acute graft-versus-host-disease in pediatric allogeneic stem cell transplantation"

### Additional File 3 – Supplementary Discussion

We found that the patients for which we observed these associations were frequently treated with vancomycin, ciprofloxacin, and especially ceftazidime at time points early post-HSCT. This is in contrast to our previous study that revealed high *Lachnospiraceae* and *Ruminococcaceae* abundances and rapid B and NK cell reconstitution in the absence of vancomycin and ciprofloxacin treatment [1]. Vancomycin and ciprofloxacin are among the broad-spectrum antibiotics that have previously been attributed a detrimental effect on the commensal microbiota, especially on *Clostridiales* [1–3]. The effect of ceftazidime however is controversially discussed. On the one hand, commensal sparing has previously been observed for cefepime, which belongs to the same antibiotic class as ceftazidime (cephalosporins) [4]. On the other hand, ceftazidime treatment could not rescue bacterial alpha diversity compared with e.g. vancomycin or ciprofloxacin treatment in a previous report [2]. In the present study, high abundances of clostridial microbiota members despite vancomycin and ciprofloxacin treatment might point to a potential beneficial effect of the additional treatment with ceftazidime. Moreover, future studies could assess if reconstitution in different NK cell subsets might differ with regards to time point and dependency on microbial composition and antimicrobial treatment.

We found that a specific *Parabacteroides* *distasonis* ASV (family *Tannerellaceae*) predicted moderate to severe aGvHD. In this context, it is interesting that our multivariate analyses indicated that a set of other *Parabacteroides* spp. ASVs was associated with high eosinophil counts from month +3 onwards. *Parabacteroides* members are providing the SCFA propionate which has previously been found to bind to the G 500 protein-coupled receptor GPR43 on eosinophils, although the exact mechanisms of GPR43-mediated immune modulation are yet to be investigated [5,6]. Interestingly, increased eosinophil numbers prior to and during aGvHD have been described before [7]. Therefore, an explanation for our prediction of moderate to severe aGvHD from high pre-HSCT *Parabacteroides* spp. abundances could involve a potential propionate-mediated activation of eosinophils, which might contribute to aGvHD. In contrast, propionate can also stimulate T_reg_ cells, which in turn prevent T_H_17-induced inflammation involved in aGvHD [8,9]. We did not observe any correlation between *Parabacteroides* members and T cell counts in our study, however T cell subsets were measured after aGvHD and thus might be altered according to the inflammatory condition itself or its treatment.

In addition, a specific *Lachnospiraceae* ASV in the gut predicted subsequent moderate to severe aGVHD when highly abundant prior to HSCT. This family has previously been reported to be reduced prior to aGvHD (but post transplantation) [10,11]. However, here we examined pre-HSCT abundances to make predictions, i.e. we did not aim at elucidating *Lachnospiraceae* abundances concurrent to aGvHD. Particular members of this bacterial family are crucial producers of the SCFA butyrate, which has been suggested to prevent aGvHD directly by improving epithelial integrity in GvHD-target tissue, and/or indirectly by inducing T_reg_ cells that reduce inflammation [9,12]. Of note, we did not observe any strong associations between bacterial abundances and inflammation (represented by CRP levels) at any body site in this study. It is of interest for future studies to reveal the exact mechanisms by which certain *Lachnospiraceae*, and specifically the predictive ASV we identified, might promote aGvHD, or whether their increase might be compensatory.

In addition to ASVs in the gut, we also identified oral and nasal cavity ASVs with pre-transplant abundances predicting subsequent aGvHD. For instance, in the oral cavity, an ASV affiliated with *Prevotella melaninogenica* strongly predicted aGvHD. This species is known to cause oral mucosal infections, and to be elevated in abundance in oral cancer [13,14]. Oral mucositis in allogeneic HSCT has been linked to elevated aGvHD risk [15,16]. It may be speculated that this could account for the observed association between aGvHD and high levels of this *Prevotella* ASV, although we did not investigate manifestations of mucositis in the present study. This highlights the need for direct evaluation of oral mucositis in this context.

Other microbial predictors of aGvHD in the oral and nasal cavity included members of the order *Actinomycetales* (*Actinomyces* spp., *Pseudopropionibacterium* *propionicum*, *Rothia* sp.). The order *Actinomycetales* comprises numerous commensals colonizing the oral and nasal cavities in healthy individuals, but can cause opportunistic infections in e.g. allo-HSCT patients [17]. It has been demonstrated that aGvHD increased the susceptibility to infections in allogeneic HSCT patients, but a predisposition to aGvHD due to a preceding infection has also been suggested for *Clostridium difficile* [18]. Whether actinomycosis might play a role in facilitating aGvHD development remains to be investigated. Interestingly, an increase of oral *Actinobacteria* in the gut at the time of neutrophil recovery has been found to be correlated with subsequent severe aGvHD [11]. One potential explanation involves the ability of *Actinomyces* spp. to drive biofilm formations, e.g. in periodontitis and during colorectal cancer [19].

Interestingly, among all ASVs significantly predicting aGvHD at any body site, we found only one ASV, namely a nasal *Rothia* sp. ASV, with high pre-HSCT abundances predicting that patients would be spared from subsequent aGvHD. *Rothia mucilaginosa* was among the species that, when present in the gut post-transplant, correlated with moderate to severe aGvHD in a previous study [11]. However, it has to date not been assessed in which way different *Rothia* species in the nasal cavity might be associated with aGvHD. This discrepancy again emphasizes the importance of high taxonomic resolution in this context, and the examination of the microbiota at different body sites.

**References**

1. Ingham AC, Kielsen K, Cilieborg MS, Lund O, Holmes S, Aarestrup FM, et al. Specific gut microbiome members are associated with distinct immune markers in pediatric allogeneic hematopoietic stem cell transplantation. Microbiome [Internet]. BioMed Central; 2019 [cited 2019 Sep 18];7:131. Available from: https://microbiomejournal.biomedcentral.com/articles/10.1186/s40168-019-0745-z

2. Weber D, Hiergeist A, Weber M, Dettmer K, Wolff D, Hahn J, et al. Detrimental effect of broad-spectrum antibiotics on intestinal microbiome diversity in patients after allogeneic stem cell transplantation: Lack of commensal sparing antibiotics. Clin Infect Dis [Internet]. 2018 [cited 2018 Sep 20]; Available from: http://www.ncbi.nlm.nih.gov/pubmed/30124813

3. Weber D, Jenq RR, Peled JU, Taur Y, Hiergeist A, Koestler J, et al. Microbiota Disruption Induced by Early Use of Broad Spectrum Antibiotics is an Independent Risk Factor of Outcome after Allogeneic Stem Cell Transplantation [Internet]. Biol. Blood Marrow Transplant. Elsevier Inc.; 2017. Available from: http://linkinghub.elsevier.com/retrieve/pii/S1083879117302756

4. Shono Y, Docampo MD, Peled JU, Perobelli SM, Velardi E, Tsai JJ, et al. Increased GVHD-related mortality with broad-spectrum antibiotic use after allogeneic hematopoietic stem cell transplantation in human patients and mice. Sci Transl Med [Internet]. 2016 [cited 2016 May 23];8:339ra71-339ra71. Available from: http://stm.sciencemag.org/content/8/339/339ra71

5. Biagi E, Zama D, Nastasi C, Consolandi C, Fiori J, Rampelli S, et al. Gut microbiota trajectory in pediatric patients undergoing hematopoietic SCT. Bone Marrow Transplant [Internet]. Nature Publishing Group; 2015 [cited 2018 Jul 2];50:992–8. Available from: http://www.nature.com/articles/bmt201516

6. Maslowski KM, Vieira AT, Ng A, Kranich J, Sierro F, Yu D, et al. Regulation of inflammatory responses by gut microbiota and chemoattractant receptor GPR43. Nature [Internet]. NIH Public Access; 2009 [cited 2018 Oct 8];461:1282–6. Available from: http://www.ncbi.nlm.nih.gov/pubmed/19865172

7. Johnsson M, Cromvik J, Johansson J-E, Wennerås C, Vaht K. Eosinophils in the blood of hematopoietic stem cell transplanted patients are activated and have different molecular marker profiles in acute and chronic graft-versus-host disease. Immunity, Inflamm Dis. 2014;2:99–113.

8. Kverka M, Zakostelska Z, Klimesova K, Sokol D, Hudcovic T, Hrncir T, et al. Oral administration of Parabacteroides distasonis antigens attenuates experimental murine colitis through modulation of immunity and microbiota composition. Clin Exp Immunol [Internet]. Wiley-Blackwell; 2011 [cited 2018 Nov 19];163:250–9. Available from: http://www.ncbi.nlm.nih.gov/pubmed/21087444

9. Arpaia N, Campbell C, Fan X, Dikiy S, Van Der Veeken J, Deroos P, et al. Metabolites produced by commensal bacteria promote peripheral regulatory T-cell generation. Nature [Internet]. Nature Publishing Group; 2013;504:451–5. Available from: http://dx.doi.org/10.1038/nature12726

10. Han L, Jin H, Zhou L, Zhang X, Fan Z, Dai M, et al. Intestinal Microbiota at Engraftment Influence Acute Graft-Versus-Host Disease via the Treg/Th17 Balance in Allo-HSCT Recipients. Front Immunol [Internet]. 2018 [cited 2018 May 17];9:669. Available from: http://www.ncbi.nlm.nih.gov/pubmed/29740427

11. Golob JL, Pergam SA, Srinivasan S, Fiedler TL, Liu C, Garcia K, et al. Stool Microbiota at Neutrophil Recovery Is Predictive for Severe Acute Graft vs Host Disease After Hematopoietic Cell Transplantation. Clin Infect Dis [Internet]. Oxford University Press; 2017 [cited 2018 Nov 23];65:1984–91. Available from: https://academic.oup.com/cid/article/65/12/1984/4085173

12. Mathewson ND, Jenq R, Mathew A V, Koenigsknecht M, Hanash A, Toubai T, et al. Gut microbiome-derived metabolites modulate intestinal epithelial cell damage and mitigate graft-versus-host disease. Nat Immunol [Internet]. NIH Public Access; 2016 [cited 2018 May 15];17:505–13. Available from: http://www.ncbi.nlm.nih.gov/pubmed/26998764

13. Olczak-Kowalczyk D, Daszkiewicz M, Krasuska-Slawińska, Dembowska-Bagińska B, Gozdowski D, Daszkiewicz P, et al. Bacteria and Candida yeasts in inflammations of the oral mucosa in children with secondary immunodeficiency. J Oral Pathol Med. 2012;41:568–76.

14. Mager DL, Haffajee AD, Devlin PM, Norris CM, Posner MR, Goodson JM. The salivary microbiota as a diagnostic indicator of oral cancer: a descriptive, non-randomized study of cancer-free and oral squamous cell carcinoma subjects. J Transl Med [Internet]. BioMed Central; 2005 [cited 2018 Dec 7];3:27. Available from: http://www.ncbi.nlm.nih.gov/pubmed/15987522

15. Correa ME, Schubert MM, Storer B, Martin PJ, Flowers MED. Correlation Between Severity of Oral Mucositis and Both Overall and Gut Acute GvHD After High Intensity Conditioning Hematopoietic Stem Cell Transplantation. Blood [Internet]. 2012 [cited 2018 Nov 25];120. Available from: http://www.bloodjournal.org/content/120/21/3057?sso-checked=true

16. Chaudhry HM, Bruce AJ, Wolf RC, Litzow MR, Hogan WJ, Patnaik MS, et al. The Incidence and Severity of Oral Mucositis among Allogeneic Hematopoietic Stem Cell Transplantation Patients: A Systematic Review. Biol Blood Marrow Transplant [Internet]. Elsevier; 2016 [cited 2018 Nov 25];22:605–16. Available from: https://www.sciencedirect.com/science/article/pii/S1083879115006394

17. Barraco F, Labussière-Wallet H, Valour F, Ducastelle-Leprêtre S, Nicolini FE, Thomas X, et al. Actinomycosis after allogeneic hematopoietic stem cell transplantation despite penicillin prophylaxis. Transpl Infect Dis. 2016;18:595–600.

18. Chakrabarti S, Lees A, Jones S, Milligan D. Clostridium difficile infection in allogeneic stem cell transplant recipients is associated with severe graft-versus-host disease and non-relapse mortality. Bone Marrow Transplant [Internet]. Nature Publishing Group; 2000 [cited 2018 Nov 26];26:871–6. Available from: http://www.nature.com/articles/1702627

19. Flynn KJ, Baxter NT, Schloss PD. Metabolic and Community Synergy of Oral Bacteria in Colorectal Cancer. mSphere [Internet]. American Society for Microbiology (ASM); 2016 [cited 2018 Nov 26];1. Available from: http://www.ncbi.nlm.nih.gov/pubmed/27303740
